# The spatial-temporal risk profiling of *Clonorchis sinensis* infection in South Korea: A geostatistical modeling study

**DOI:** 10.1101/2022.04.03.22273349

**Authors:** Hai-Yan Xiao, Jong-Yil Chai, Yue-Yi Fang, Ying-Si Lai

## Abstract

Clonorchiasis is one of the major parasitic diseases in South Korea. Spatially explicit estimates of the infection risk are important for control and intervention. We did a systematic review of collected prevalence data on *Clonorchis sinensis* infection in South Korea. Data of potential influencing factors (e.g., environmental and social-economic factors) was obtained from open-access databases. Bivariate Bayesian geostatistical joint modeling approaches were applied to analyze disease data, within a logit regression in combination of potential influencing factors and spatial-temporal random effects. We identified surveys of *C. sinensis* infection done at 1362 unique locations, and presented the first spatial-temporal risk maps at high spatial resolution (5×5km) in South Korea. High infection risk areas shrunk significantly from 1970 to 2017. The overall risk decreased since the start of the national deworming program in 1969, and then slightly increased since the year 1995 when the program suspended, and maintained stable since 2005 when the Clonorchiasis Eradication Program begun. The population-weighted prevalence was estimated as 3.87% (95% BCI: 3.04-4.82%) in 2017, accounting to 1.92 (95% BCI: 1.51-2.40) million infected people. Although the prevalence over the country has been low, *C. sinensis* infection was still endemic in areas of eastern and southern regions, particularly the five major river basins. We also defined factors significantly correlated, such as, distance to the nearest open water bodies, annual precipitation, and land surface temperature at night. All findings above provide important information on spatial-targeting control and preventive strategies of *C. sinensis* infection in South Korea.

## Introduction

Clonorchiasis, caused by infection with *Clonorchis sinensis*, is one of the most important food-borne trematodiases in humans (***Na et al., 2020; Qian et al., 2016***). Ingestion of raw or undercooked freshwater fish infected with *C. sinensis* metacercaria is the main way for humans to be infected (***Qian et al., 2020; Qian and Zhou, 2021***). The parasite can damage the liver and biliary systems, which may lead to cholelithiasis, cholangitis, or even fatal cholangiocarcinoma (***Na et al., 2020; Qian et al., 2012; Qian et al., 2016***). Preventive chemotherapy, information, education, and communication (IEC), environmental modification and the possible combinations of the three are considered as major control measures of the disease (***Huang et al., 2020; Qian et al., 2016***). The infection is endemic mainly in Asia, and countries reporting the disease including China, South Korea, Vietnam and Russia (***Fürst et al., 2012; Qian and Zhou, 2021; Rim, 2005***). Particularly, clonorchiasis is one of the major parasitic diseases in South Korea, the total medical expense of which was approximately 222 million won in 2018, ranking the fourth among all parasitic diseases in the country (***Kim et al., 2019***).

According to the results of the national surveys, the raw prevalence of *C. sinensis* infection in South Korea were 4.6%, 1.8%, 2.6%, 2.7%, 2.2%, 1.4%, 2.9% and 1.9% in the year 1971, 1976, 1981, 1986, 1992, 1997, 2004 and 2012, respectively (***The Ministry of Health and Social Affairs and The Korean Association of parasite Eradication, 1971; The Ministry of Health and Social Affairs and The Korean Association of parasite Eradication, 1976; The Ministry of Health and Social Affairs and The Korea Association of Parasite Eradication, 1982; The Ministry of Health and Social Affairs and The Korea Association of Health, 1986; The Ministry of Health and Social Affairs and The Korea Association of Health, 1993; The Ministry of Health and Welfare and Korea Association of Health Promotion, 1997; Korea Association of Health Promotion, 2004; Korea Centers for Disease Control and Prevention and Korea National Institute of Health, 2013***), suggesting a relatively low overall prevalence of the disease in the country.

However, the distribution of the infection shows diversity in space and time. Residents living in river basins have higher prevalence compared to those living elsewhere (***Lee et al., 2020; Shin et al., 2017***). Besides, as shown in surveys of *C. sinensis* conducted in the five major river basins of South Korea, there was an overall decline in prevalence over years, however, the prevalence in different river basins differed considerably, with different degrees and trends of decline (***Chai, 1990; Cho et al., 2016; Jeong et al., 2016; June, 2009; June et al., 2013; Kim et al., 2016; Kim et al., 2010; Shin et al., 2020***). Since the national survey in 2012 (***Korea Centers for Disease Control and Prevention and Korea National Institute of Health, 2013***), surveys on *C. sinensis* infection were mainly focused on the endemic areas around the five major river basins (***Jeong et al., 2016; Kim et al., 2016; Lee et al., 2020; Shin et al., 2017***). Understanding the spatial-temporal distribution of *C. sinensis* infection at high resolution is of great importance for spatial-targeting control and preventive strategies in the country. However, to our knowledge, high-resolution spatial-temporal risk maps of *C. sinensis* infection haven’t been produced in South Korea, and simple statistical description of the historical survey data is not enough to obtain this information.

Bayesian geostatistical modeling is one of the most rigorous approaches to produce high-resolution disease risk maps (***Karagiannis-Voules et al., 2015***). It has been applied in various studies to food-borne trematodiases or other neglected tropical diseases in many regions around the world, such as clonorchiasis in China, opisthorchiasis in Southeast Asia, schistosomiasis in Africa and soil-transmitted helminthiasis in South Asia (***Lai et al., 2015; Lai et al., 2019; Lai et al., 2017; Zhao et al., 2021***). This approach usually models point-referenced disease data with potential risk factors (e.g., socioeconomic and environmental factors) and spatial-temporal random effects, thus estimates disease risk in areas without observed (***Gelfand and Banerjee, 2017; Lai et al., 2015***). Recently, more advanced Bayesian geostatistical models were developed to deal with data heterogeneities, such as jointly analyzing diseases data at both point- and areal levels (***Moraga et al., 2017; Smith et al., 2008***), and joint modeling of disease data which is partially missed the total number examined (***Zhao et al., 2021***)

In this study, we aimed to understand the spatial-temporal distribution of *C. sinensis* infection in South Korea. We collected available disease survey data and data of potential influenced factors, based on which, a Bayesian geostatistical joint model was built and yearly risk maps of *C. sinensis* infection in South Korea at 5×5 km^2^ were produced.

## Results

### Data summaries

The data selection flow diagram is presented in Figure 1. A total of 2448 records were identified through databases, while an additional 4 records were stemmed from government reports 112 records remained according to the inclusion and exclusion criteria, resulting in 57 surveys at 15 ADM1-level divisions, 226 surveys at 185 ADM2-level divisions, and 1,404 surveys at 1184 point-referenced locations. According to the quality assessment checklist, 96.43% of the included records showed good quality, with scores no less than 6 (Figure 2, Supplementary Table 1). Figure 4 shows the geographic distribution of locations and observed prevalence of *C. sinensis* infection. Table 1 provides an overview of survey characteristics, categorized by different periods (i.e., 1970-1976, 1977-1984, 1985-1992, 1993-2000, 2001-2008, and 2009-2017). Large proportion of surveys (23.59%) were conducted during the period 2001 to 2008 and the less proportion (8.36%) during 1970 to 1976. Around 96.32% of surveys are community based. Kato-Katz method was the most commonly used diagnostic method (76.11%), followed by Formalin-ether sedimentation technique (FE). The mean prevalence, calculated directly from all survey data, was 7.43%.

**Table 1.** Overview of *C. sinensis* infection survey data in South Korea.

| Year of survey | 1970-1976 | 1977-1984 | 1985-1992 | 1993-2000 | 2001-2008 | 2009-2017 | Total |
| --- | --- | --- | --- | --- | --- | --- | --- |
| Number of articles | 14 | 34 | 16 | 22 | 21 | 9 | 112 |
| Number of surveys/locations<sup>a</sup> | 141/140 | 247/232 | 279/261 | 295/277 | 398/395 | 327/292 | 1687/1362 |
| Location type<sup>a</sup> |  |  |  |  |  |  |  |
| ADM1 | 11/10 | 8/4 | 28/14 | 0/0 | 1/1 | 9/9 | 57/15 |
| ADM2 | 7/7 | 14/12 | 9/9 | 6/6 | 10/9 | 180/179 | 226/185 |
| Points | 123/123 | 225/216 | 242/238 | 289/271 | 387/385 | 138/104 | 1404/1184 |
| Survey type<sup>a</sup> |  |  |  |  |  |  |  |
| Community | 139/138 | 220/207 | 275/259 | 278/263 | 388/385 | 325/291 | 1625/1314 |
| School | 2/2 | 27/27 | 4/4 | 17/17 | 10/10 | 2/2 | 62/59 |
| Stool test<sup>a</sup> |  |  |  |  |  |  |  |
| Kato-Katz | 109/108 | 209/199 | 245/230 | 214/214 | 295/295 | 212/211 | 1284/1084 |
| FE<sup>b</sup> | 32/32 | 27/26 | 12/11 | 39/37 | 91/89 | 115/81 | 316/266 |
| FE and Kato-Katz<sup>c</sup> | 0/0 | 11/10 | 22/22 | 42/40 | 12/12 | 0/0 | 87/81 |
| Average prevalence (%) | 8.12 | 10.85 | 6.86 | 6.91 | 7.50 | 5.41 | 7.43 |

<sup>a</sup>Presented as surveys/locations.<sup>b</sup>FE: formalin-ether concentration technique.<sup>c</sup>Reporting results based on both FE and Kato-Katz.

**Figure 1.**
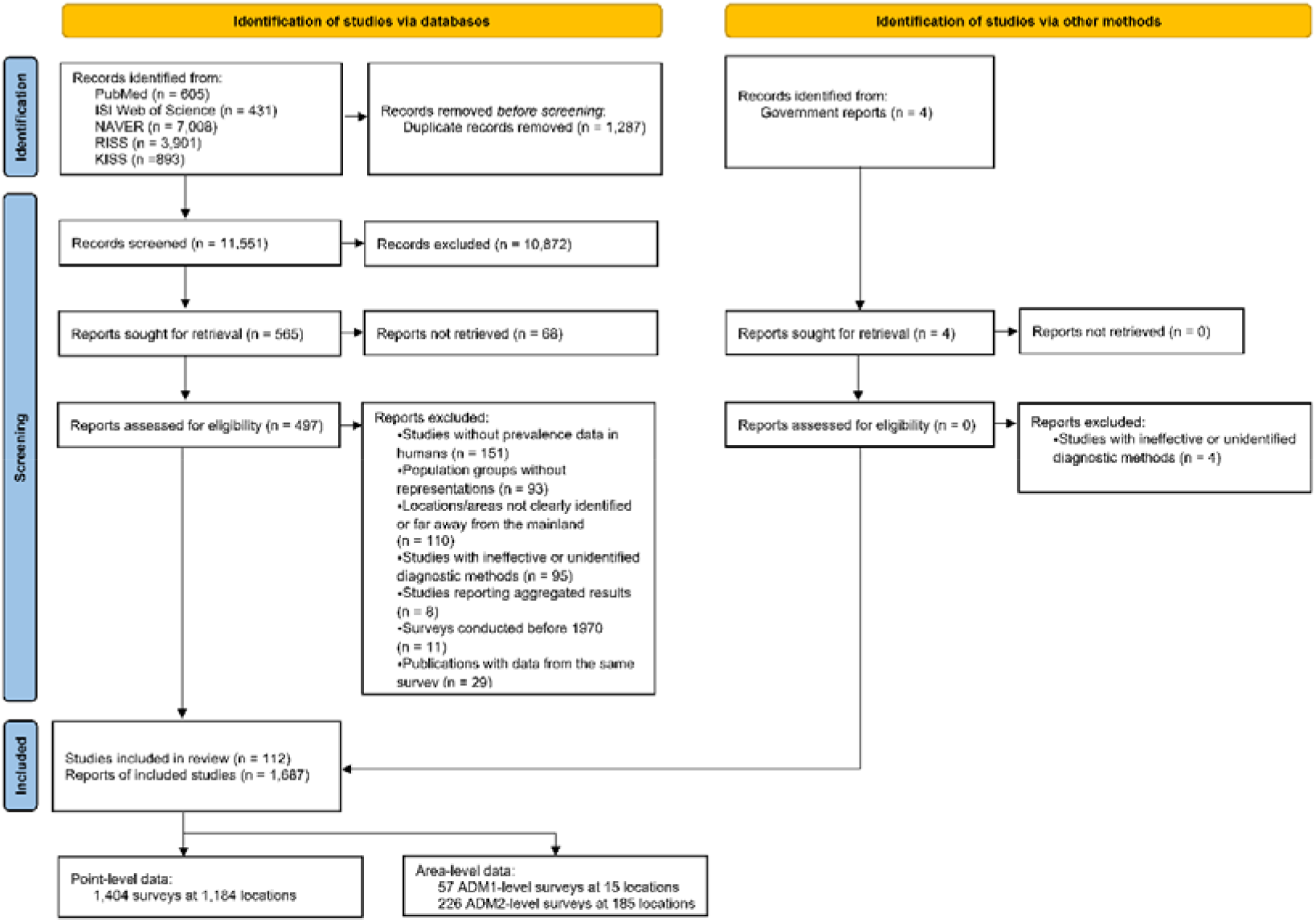
Data selection flow chart.

**Figure 2.**
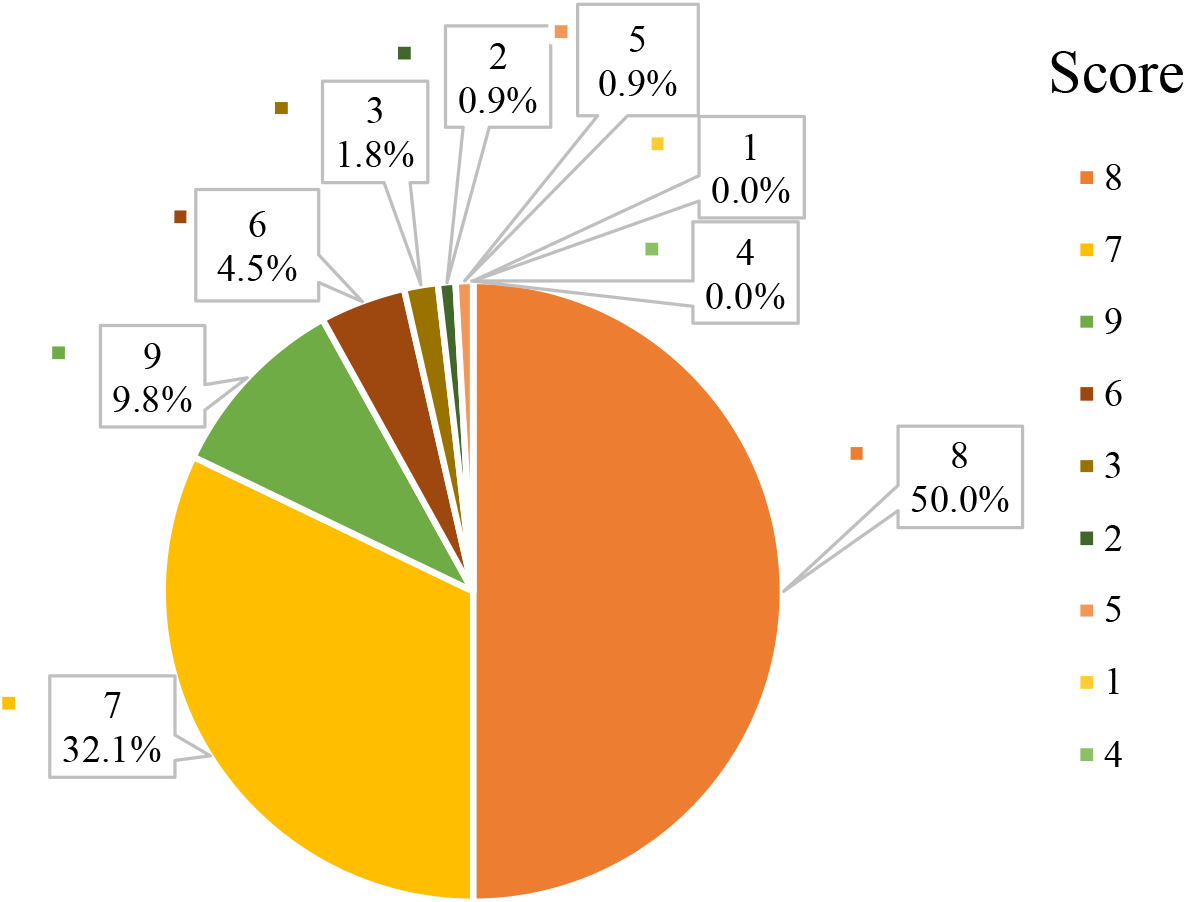
The quality assessment of eligible studies. Each piece represents the number and proportion of studies under the corresponding quality score.

### Modeling fitting

After variable selection, a total of seven variables (i.e., human influence index, annual precipitation, distance to the nearest open water bodies, land surface temperature at night, soil moisture, normalized difference vegetation index and travel time to the nearest big city) were selected for the final geostatistical model (Table 2). The infection risk was 4.30 (95% BCI: 3.32-5.60) times in the community as much as that in school-aged children. The risk of *C. sinensis* infection was lower in people living in urban areas compared to those living in rural areas. A negative association was found for the infection risk with the distance to the nearest open water bodies and annual precipitation, whereas a positive relationship was identified with land surface temperature at night and normalized difference vegetation index.

**Table 2.** Posterior summaries of model parameters for *C. sinensis* infection.

| Variable | Estimated median | OR | Prob (%)<sup>a</sup> |
| --- | --- | --- | --- |

|  | (95% BCI) |  |  |
| --- | --- | --- | --- |
| Intercept | -5.41(-5.98, -4.85)<sup>b</sup> | 0.005(0.003, 0.01) | <0.01 |
| Survey type (Community)<sup>c</sup> |  |  |  |
| School | 1.46(1.20, 1.72)<sup>b</sup> | 4.30(3.32, 5.60) | >99.99 |
| Urban extents (Rural)<sup>c</sup> |  |  |  |
| Urban | -0.53(-0.83, -0.24)<sup>b</sup> | 0.59(0.44, 0.79) | <0.01 |
| Land surface temperature in the daytime ($\leq 17.0^{\circ}\text{C}$)<sup>c</sup> | | | |
| 17.0-18.5 | 0.13(-0.12, 0.39) | 1.14(0.88, 1.47) | 80.40 |
| > 18.5 | 0.24(-0.12, 0.61) | 1.27(0.88, 1.84) | 89.20 |
| Annual precipitation ($\leq 1266\text{mm}$)<sup>c</sup> | | | |
| 1263-1366 | -0.46(-0.82, -0.09)<sup>b</sup> | 0.63(0.44, 0.91) | 0.40 |
| > 1366 | -0.51(-0.98, -0.04)<sup>b</sup> | 0.60(0.38, 0.96) | 1.20 |
| Elevation ($\leq 56\text{m}$)<sup>c</sup> | | | |
| 56-129 | 0.04(-0.19, 0.28) | 1.04(0.82, 1.32) | 53.40 |
| > 129 | -0.18(-0.50, 0.15) | 0.84(0.61, 1.16) | 20.00 |
| Distance to the nearest open water bodies (km) | -0.51(-0.65, -0.38)<sup>b</sup> | 0.60(0.52, 0.69) | <0.01 |
| Land surface temperature at night (□) | 0.19(0.003, 0.39)<sup>b</sup> | 1.21(1.00, 1.47) | 96.60 |
| Nighttime light | -0.06(-0.30, 0.18) | 0.94(0.74, 1.19) | 59.40 |
| Normalized difference vegetation index | 0.24(0.06, 0.43)<sup>b</sup> | 1.28(1.06, 1.54) | 99.60 |
| Range (km) | 45.60(38.60, 53.66) | - |  |
| Spatial variance ($\sigma_{\phi}^2$) | 7.61(6.33, 9.19) | - | |
| Non-spatial variance ($\sigma_{nonsp}^2$) | 0.64(0.50, 0.81) | - | |
| Temporal correlation coefficient ($\rho$) | 0.04(-0.13, 0.20) | - | |
| Variance of beta-likelihood ($\sigma_{\beta}^2$) | 0.01(0.01, 0.02) | - | |

<sup>a</sup>Posterior probability of OR > 1.
<sup>b</sup>Important effect based on 95% Bayesian credible interval (BCI).
<sup>c</sup>In brackets, baseline values are reported.

### Validation of model

Model validation showed that the model was able to correctly estimate 79.60% of locations within the 95% BCI, and the AUC of ROC was 0.86, suggesting a reasonable capacity of prediction accuracy of our model. The ME, MAE and MSE were 0.88%, 5.00%, and 1.06%, respectively, in the final Bayesian model.

### Risk maps and estimates of number of people infected

Figure 4 presents the model-based estimated risk maps of *C. sinensis* infection in selected years (i.e., 1970, 1977, 1985, 1993, 2001, 2009 and 2017) in South Korea. Overall, high infection risk (with prevalence>20%) areas shrunk across the study period, but temporal variance was shown in different areas. There was an obvious decrease of infection risk in the central region (e.g., Chungcheongbuk-do and the western part of Gyeongsangbuk-do), which was once the highest-risk region. The southern areas (e.g., Gyeongsangnam-do, Jeollanam-do, Gwangju Gwangyeoksi, Ulsan Gwangyeoksi, and Busan Gwangyeoksi) firstly showed a trend down from 1970 to 1993, then a moderate increase until 2001, which was followed by a gradual decline since then. The infection risk of *C. sinensis* in the eastern region (e.g., The central and eastern parts of Gyeongsangbuk-do) was estimated to be on a downward trend until around 2009, followed by a slight upward trend, and several areas remained moderate risk (with prevalence between 5% and 20%) in recent years. Most areas in the western and northern regions (e.g., Chungcheongnam-do, Gangwon-do, and Gyeonggi-do) showed stably low infection risk (<5%). Particularly, the moderate to high infection risk areas distributed mainly along river basins. High estimation uncertainty was presented in several areas of the central, the southern and the eastern regions of the country (Figure 5).

**Figure 3.**
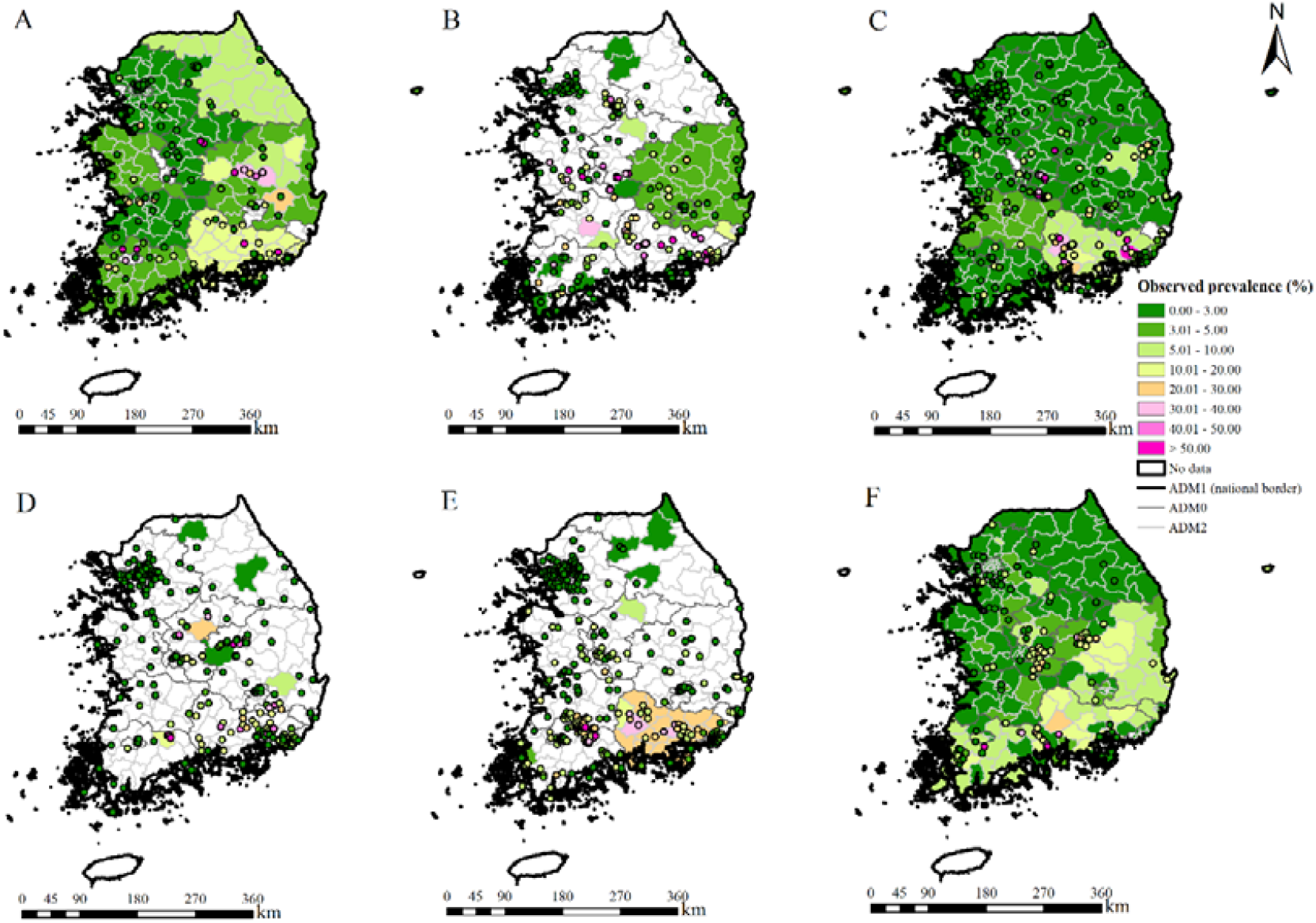
Survey locations and observed prevalence of *C. sinensis* infection in South Korea. (A) 1970–1976, (B) 1977–1984, (C) 1985–1992, (D) 1993–2000, (E) 2001–2008, (F) 2009–2017.

**Figure 4.**
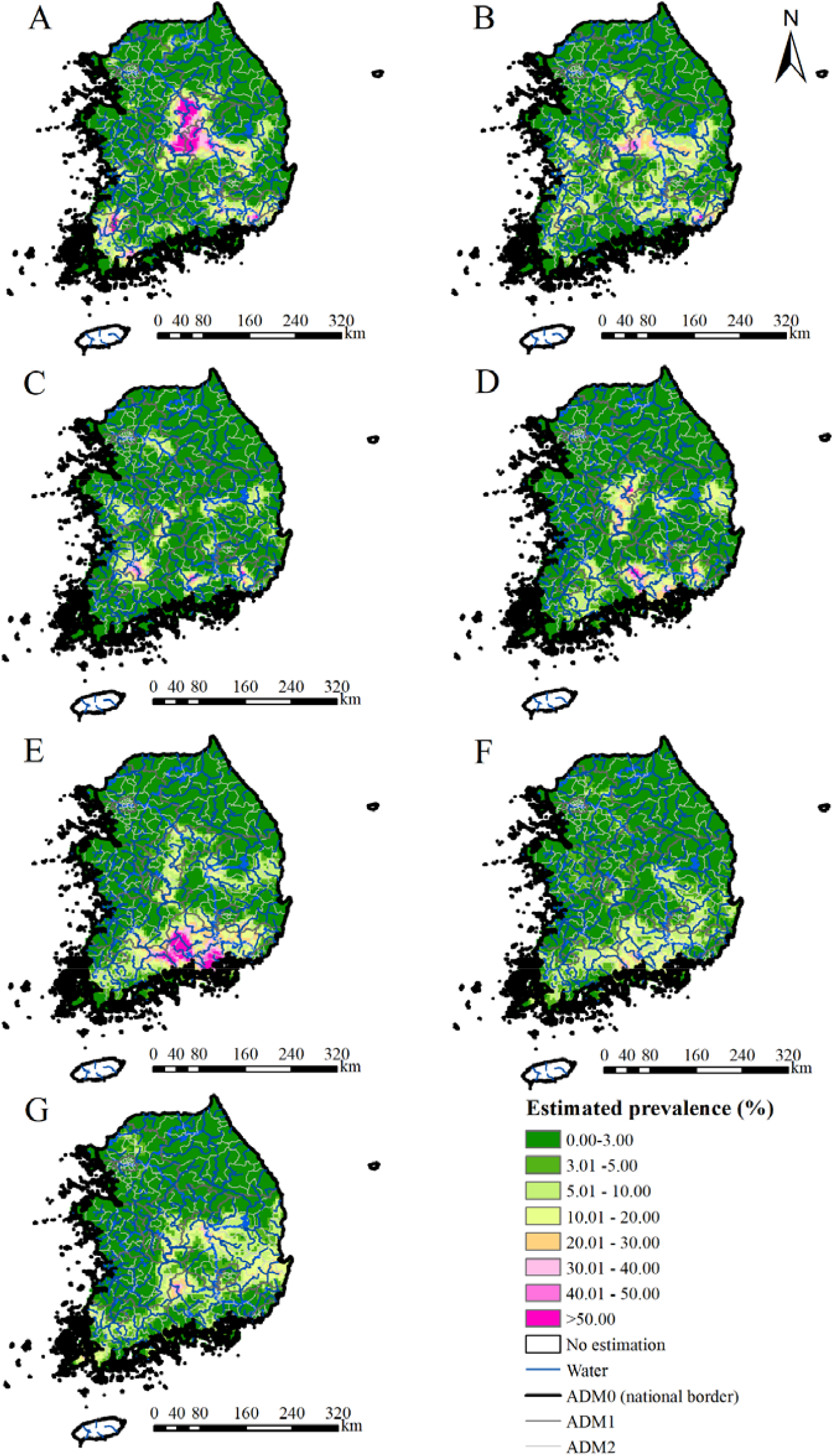
Model-based estimated risk maps of *C. sinensis* infection across South Korea in selected years. Estimated prevalence was based on the median of the posterior estimated distribution of infection risk in (A) 1970, (B) 1977, (C) 1985, (D) 1993, (E) 2001, (F) 2009, and(G) 2017.

**Figure 5.**
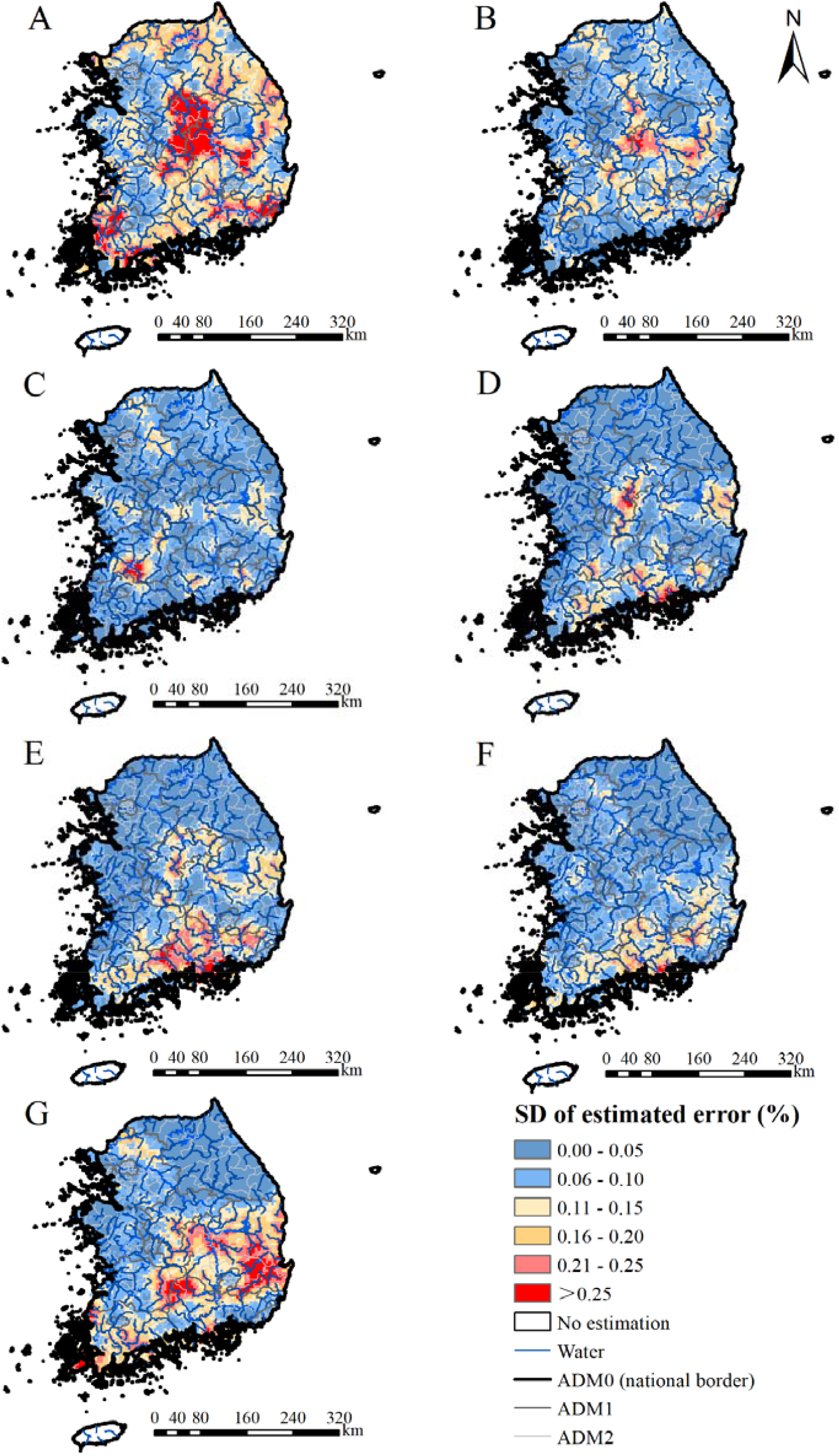
The estimation uncertainty in selected years. Results were based on the standard. deviation of the posterior estimated distribution of infection risk in (A) 1970, (B) 1977, (C) 1985, (D) 1992, (E) 2001, (F) 2009, and (G) 2017.

**Figure 6.**
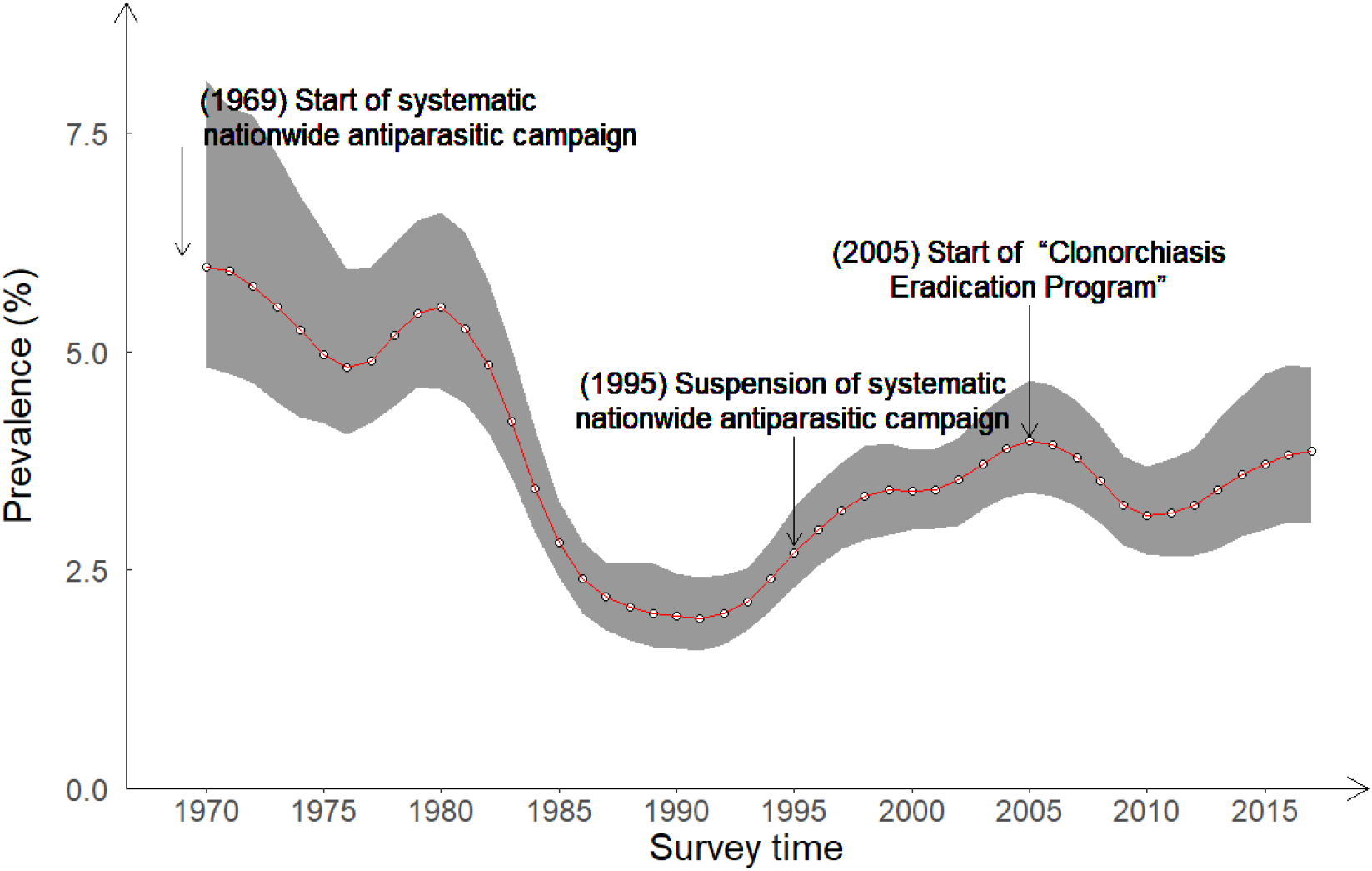
Trends in estimated prevalence of *C. sinensis* infection in South Korea.

The overall estimated infection prevalence across the country showed an obvious decrease trend until around 1991 (with the lowest estimated prevalence as 1.95%, 95% BCI 1.58-2.41%), followed by a slight increase until around 2005, and maintained stable ever since (Figure 6). In 2017, the population-adjusted estimated prevalence and the number of individuals infected with *C. sinensis* were 3.87% (95% BCI: 3.04-4.82%) and 1.92 million (95% BCI: 1.51-2.40 million), respectively.

## Discussion

To our knowledge, we presented the first spatial-temporal risk maps of *C. sinensis* infection at spatial high-resolution (5×5 km^2^) in South Korea, using rigorous Bayesian geostatistical joint modelling approach with available disease survey data and potential influencing predictors. Our study identifies high risk areas in different periods and presents the trend of infection risk with time in different areas, which provides an important reference for understanding the risk of *C. sinensis* infection in South Korea in the last 50 years and may provide valuable information for spatially targeting intervention.

Our findings on an obvious decline of the overall infection risk from 1970 to 1995 across the country suggested the effective control of *C. sinensis* infection in the period, which may be largely attributed to the nationwide biannual school-based mass fecal screening and anthelmintic-administration program implemented by the Korea Association of Health Promotion from 1969 to 1995 (***Hong and Yong, 2020; Park, 2008***). There was an increase of the risk from 1996 to 2004, probably due to the fact that the nationwide antiparasitic campaign was suspended and no large-scale control and prevention measures carried out in this period, with the exception of some small-scale interventions (***Lee et al., 2002***). For effective control of *C. sinensis* infection, the Korea Centers for Disease Control and Preventions launched the Clonorchiasis Eradication Program (CEP) in 2005, with the goal to reduce the infection rate by diagnosing, treating, and educating residents in endemic areas and the elderly (***Hong and Yong, 2020; Jo, 2013; Park, 2008***). Of note, CEP seemed effective in certain sense, at least the overall infection risk did not increase but remain stable since 2005. In practice, the CEP started in different times across different areas with different intensity of implementation, which may lead to different local trends of infection risk (***Jeong et al., 2017***).

Compared with results from the eight national surveys, we estimated comparable or slightly higher *C. sinensis* infection prevalence (Supplementary Table 2). The national surveys mainly reported raw overall prevalence by calculating the proportion of positive individuals among all examined ones (***Korea Centers for Disease Control and Prevention and Korea National Institute of Health, 2013***), which may be influenced by sampling framework (***Mitani et al., 2021; Zhao et al., 2021***). Differently, our estimates relied on rigorous Bayesian geostatistical modeling of all available geo-referenced disease survey data along with important environmental and socioeconomic predictors, thus was able to take into account high-resolution spatial heterogeneous of both disease infection risk and population density across the country when compiling country-level prevalence (***Lai et al., 2019; Zhao et al., 2021***). We did not provide estimates of infection risk for islands Jeju-do and Ulleung-gun, as sparse geo-referenced data were obtained, and they are quite a distance away from the continent of South Korea. It is improper to combine island data and continent data for geostatistical modeling, due to spatial discontinuity (***Santafé et al., 2021***). Though, previous surveys suggested very low prevalence of *C. sinensis* infection in the two islands (0%∼0.58% in the previous national surveys for Jeju-do from 1971 to 2012, and 0.4% in a survey in 1973 for Ulleung-gun) (***The Ministry of Health and Social Affairs and The Korean Association of parasite Eradication, 1971; The Ministry of Health and Social Affairs and The Korean Association of parasite Eradication, 1976; The Ministry of Health and Social Affairs and The Korea Association of Parasite Eradication, 1982; The Ministry of Health and Social Affairs and The Korea Association of Health, 1986; The Ministry of Health and Social Affairs and The Korea Association of Health, 1993; The Ministry of Health and Welfare and Korea Association of Health Promotion, 1997; Korea Association of Health Promotion, 2004; Korea Centers for Disease Control and Prevention and Korea National Institute of Health, 2013; Chai, 1990; Cho et al., 1973; Kim et al., 1971; Se et al., 1972; Shim et al., 1982***).

We identified several socioeconomic and environmental predictors significantly related with *C. sinensis* infection, which may provide insights in prevention and control of the disease. We found that community-based surveys were tended to report higher prevalence than that in schoolchildren, in line with the results of previous studies (***Lee et al., 1994a, 1994b; Min et al., 2002; Zhao et al., 2021***). Some studies suggested that the youth eat raw fish far less frequently than the elderly, resulting in a generally lower prevalence among the youth (***June et al., 2013; Lee et al., 2020***). The prevention *C. sinensis* infection may more focus on the elderly (***June et al., 2013; Lee et al., 2020***), though should not ignore the youth. Our results showed that the infection risk was higher in rural areas comparing to that in urban areas, similar with findings from other studies (***Kim, 1983; Moon et al., 1981; Song et al., 1981***). People living in rural areas may be weaker in personal hygiene awareness and with poor sanitation conditions, probably due to less education or economic disadvantage (***Kim, 1983; Song et al., 1981***). In addition, the culturally rooted habits of eating raw or insufficiently cooked freshwater fish seemed more common in rural (***Moon et al., 1981***). This suggested that the prevention *C. sinensis* infection in rural areas should not be neglected. We found that distance to the nearest water bodies had a negative effect with *C. sinensis* infection, consistent with knowledge that inhabitants living closer to waterbodies had more chance of freshwater fish consumption (***Jeong et al., 2017; Lai et al., 2017***). The above finding suggested the rural elderly population in the river basins should be prioritized for effective control and prevention of *C. sinensis* infection. In addition, we found that environmental factors, such as land surface temperature at night, normalized difference vegetation index, and precipitation, were associated with infection risk, possibly due to the fact that environmental factors may influence the survival and reproduction of fish, snails, caecilians, etc., and thus affect the risk in the corresponding areas (***Lai et al., 2017***).

Frankly, there are several limitations in our study. As we collected all available survey data through a systematic review, the survey locations seemed unevenly distributed across the study region, that is a greater number of surveys were from both the high and the low endemic areas while fewer surveys were from moderate endemic areas. Thus, preferential sampling may exist (***Zhao et al., 2021***). However, current approaches handling preferential sampling issue targets just on “one-way preference”, that is more samples were draw from higher or lower risk areas, different from our “two-way” situation that more samples were from both higher and lower risk areas. Thus, these approaches were not proper to solve the current issue (***Krainski et al., 2019***). Nevertheless, our model performance was good, with prediction accuracy 79.60% within 95% BCI and AUC under ROC 0.86, suggesting that our results were reliable. We are currently developing models handling the “two-way” preferential sampling issue. Secondly, age and sex were found significantly associated with *C. sinensis* infection risk (***Park, 2007; Park et al., 2014***). However, as most surveys didn’t report sex- and age-specific infection prevalence, we were not able to consider these factors in the modeling analysis. Nevertheless, we considered the survey type (i.e., community- and school-based) in the models, thus partially taking into account the potential influencing effect of age on infection risk. Thirdly, we included surveys with different diagnostic techniques (with Kato-Katz and FE the major techniques adopted). Hong et al’s study suggested that there was no significant difference in diagnostic ability between Kato-Katz and FE (***Hong et al., 2003***), while Li et al’s results showed less sensitivity of Kato-Katz than FE (***Cho et al., 1969; Uga et al., 2010***). Regarding the diagnostic abilities of different methods, the results of those studies above were inconsistent. Due to lack of more powerful studies on the diagnostic abilities of different techniques, we didn’t take into account the effect of different diagnostic methods in the final model. Nevertheless, sensitivity analysis was conducted to evaluate the effect. Similar estimated posterior distribution of parameters and consistent patterns of risk maps (Supplementary Table 3, Supplementary Figure 1) were found between the model without considering the diagnostic techniques and that with adjusting the sensitivity of surveys according to Li et al’s study (***Li et al., 1998***), suggesting that our final results were reliable. Additionally, fecal examination, especially Kato-Katz, has been widely used in mass examinations in South Korea (***Korea Centers for Disease Control and Prevention and Korea National Institute of Health, 2013; Jeong et al., 2016***), but it is difficult for inexperienced researchers to identify the eggs of *C. sinensis, Metagonimus yokogawai, Metagonimus miyatai*, and *Metagonimus takahashii* by microbiological methods alone without using molecular diagnostic methods such as PCR (***Jeon et al., 2012; Xu et al., 2019***). The high prevalence of *C. sinensis* infection in Hadong-gun and Gokseong-gun, two countys with high prevalence of *M. yokogawai* infection previously (***Bahk et al., 2018; Chai et al., 2015; Kim et al., 1979***), may be caused by the absence of differential diagnosis. In addition, as seven surveys only reported the intervals of prevalence instead of the exact observed values, there may be a potential bias resulted from assigning the midpoint values of the intervals as observations (***Korea Centers for Disease Control and Prevention and Korea National Institute of Health, 2013; Cho et al., 2016; Ju et al., 2005***). Nevertheless, the sensitivity analysis suggested that this effect was ignorable, as our final estimated posterior distribution of parameters and patterns of risk maps were similar with those assigning the lower or upper bounds of the intervals as observations in these surveys (Supplementary Table 4, Supplementary Figure 2). In conclusion, we present the first temporal risk maps of *C. sinensis* infection at high-resolution across 50 years in South Korea. The overall trend in the prevalence suggested the national deworming programs from 1969 and the Clonorchiasis Eradication Program from 2005 were effective. Even though low average level of infection risk in the country, the risk maps suggested that several areas in the eastern and southern regions especially in the five major river basins were still in moderate to high infection risk, which should be prioritized for control and intervention. All findings provide important information on spatial-targeting control and preventive strategies on *C. sinensis* infection in South Korea.

## Methods

### Ethics statement

This work was based on survey data of clonorchiasis, which was derived from peer-reviewed published literatures. Statements of ethics approval were included in the original sources. All data in this study were aggregated at point- or area-level and did not contain any identifiable information at the individual or household levels. So, there are no specific ethical issues warranted.

### Disease data

We did a systematic review to collect data related to prevalence of *C. sinensis* infection in South Korea (registered in the International Prospective Register of Systematic Reviews No. CRD42021234803), and reported it following the PRISMA guidelines (Supplementary File 1, Supplementary File 2) (***Page et al., 2021***). Two general databases (PubMed and ISI Web of Science) and three South Korea databases (NAVER, KISS and RISS) were searched from inception to June 4, 2020, with search terms “(Liver fluke* OR Clonorchi*) AND Korea” and ““간흡충“ OR ”간디스토마; OR “*Clonorchis sinensis*” OR “Clonorchiasis””, respectively. There were no restrictions on publication time, language and study design in our search strategy. Additionally, we considered other potential literatures, such as reports from governments or Ministry of Health, documents from research groups, relevant books and theses. The literatures obtained from different sources was pooled together and duplicated ones were removed.

A protocol is listed in Supplementary Figure 3, with clear criteria for inclusion, exclusion and extraction of data. In brief, we included prevalence-related surveys (i.e., with information on number of examined and number of positive, or information on prevalence) conducted from 1970 onwards, reporting at provincial level and below, such as administrative divisions of level one (ADM1: province, special city, etc.), level two (ADM2: city, county, etc.) and at point-level (village, school, etc.). Studies were excluded if they were in-vitro investigations, or absence of human studies, or with specific study designs (e.g., case-control studies) or specific population groups (e.g., patients), or with study locations/areas not clearly identified, or with ineffective or unidentified diagnostic methods (e.g., direct smear, salt water flotation, serum diagnostics, intradermal test, etc.), or reporting aggregated results of both community and school populations, or with small sample size (less than 10).

Two independent reviewers assessed the quality of each included literature using an adapted nine-point quality assessment checklist (***Zhao et al., 2021***) (Supplementary File 3). We followed the GATHER checklist (Supplementary File 4) for the data extraction All included data were georeferenced by Google Maps (https://www.google.com/maps/) and entered into a database with detailed information (e.g., literature information, survey information, location information and disease-related data). For surveys reported intervals of prevalence instead of exact observed values, we assigned the midpoints of the intervals as the observed prevalence. If studies adopted multiple diagnostic methods and reported the observed prevalence accordingly, the one with the most sensitive diagnostic method was extracted. If survey year was missing, we approximated it with the public year minus three.

### Socioeconomic, environmental and demographic data

Potential influencing data, including the socioeconomic data (i.e., nighttime light, human influence index, urban extents, and travel time to the nearest big city) and environmental data (i.e., distance to the nearest open water bodies, soil moisture, annual precipitation, elevation, land cover, land surface temperature in the daytime and at night, and normalized difference vegetation index), and the demographic data were obtained from open accessible data sources, as listed in Supplementary Table 5 and Supplementary Figure 4. Data were integrated to a regular grid of 5×5 km^2^ spatial resolution. To simplify the variable selection process, similar classes of land cover were re-grouped into five categories, that is (i) forests, (ii) grasslands and shrub, (iii) wetlands and water bodies, (iv) croplands, and (v) buildings and barren land. As the demographic data at high-spatial resolution were only available from 2000 onwards, population for previous years were estimated based on the available population data of the nearest year and the population growth rates accessed from the WHO official website (https://population.un.org/wpp/Download/Standard/Population/) with formula P_m_ = P_n_ * e^(m-n)*Tate^ where P_m_ and P_m_ are the population counts in year n and year m, respectively.

### Statistical analysis

We transformed multi-categorical predictors into dummy variables and standardized continuous ones with mean zero and standard deviation one. To avoid collinearity, Pearson’s correlation between each pair of continuous predictors was calculated. If correlation coefficients greater than 0.8, we chose the ones more meaningful or with better data quality among pairs. To build a parsimonious model, Bayesian variable selection was adopted to find the best set of predictors. First, to identify the best functional form (continuous or categorical) of continuous predictors, we converted them to three-level categorical ones according to preliminary, exploratory, graphical analysis (***Lai et al., 2017***). Bayesian geostatistical models was built with either form as the independent variable. The one with the lowest log score was selected as the best functional form. Second, we considered all possible combinations of potential predictors with their best functional forms, and fitted Bayesian geostatistical models with each combination. The one with the lowest log score was selected as the final model. To be noted, survey type (community- or school-based) was kept in all combinations, according to the previous studies suggesting infection risk different between community population and school-aged children (***Lee et al., 1993; Lee et al., 1994a, 1994b; Min et al., 2002; Song et al., 1983***).

We adopted a bivariate Bayesian geostatistical joint modeling approach proposed by Zhao Tingting and colleagues, to analyze the point- and areal level survey data together (***Moraga et al., 2017; Utazi et al., 2019***), and include disease survey data reporting both the number of examined and positive, and those only reporting prevalence (***Zhao et al., 2021***). In brief, we assumed a binomial distribution for disease data reported both the number of examined and positive, and a beta distribution for those with only prevalence reported. A logit scale of prevalence was modelled with a linear combination of fixed effect predictors, spatial-temporal random effect and exchangeable non-spatial random effect. For areal level survey data, the average values of predictors and the average spatial-temporal random effects of pixels within the corresponding areas were assigned. We assumed the spatial-temporal random effect arising from a zero-mean Gaussian distribution, with the covariance matrix the Kronecker product of a spatial matrix and a temporal one. The former was assumed following a stationary Matérn covariance function and the latter with auto-regressive order 1 (AR1). In addition, regular temporal knots were set and latent spatial-temporal random fields were approximated by employing the B-spline basis function, to reduce the computational burden. The models were built under a Bayesian framework and INLA-SPDE approach was used for model fitting (***Cameletti et al., 2013; Krainski et al., 2019***). More details were shown in Supplementary File 5.

We used the 5-fold out-of-sample cross-validation approach for model validation. To evaluate the model performance, we calculated mean error (ME), mean absolute error (MAE), mean square error (MSE), the percentage of observations covered by 95% Bayesian credible intervals (BCIs) of posterior estimated prevalence, and the area under the receiver-operating characteristic (ROC) curve (AUC) (***Brooker et al., 2001; Lai et al., 2015; Zhao et al., 2021***). More details on the model validation were shown in Supplementary File 6. In addition, sensitivity analysis was conducted to assess the effect of using the midpoint values of intervals as the observed prevalence, and the effect of different diagnostic methods (Supplementary File 7, Supplementary File 8).

A regular grid of 5×5 km^2^ spatial resolution was overlay across South Korea, resulting in 5,946 pixels. The infection risk for each pixel each year from 1970-2017 was estimated using Bayesian kriging. All the statistical process was done in R (version 4.0.4) and risk maps were produced using ArcGIS (version 10.2). The country-level infection prevalence was calculated with population-weighted pixel-level infection risk.

## Data Availability

All web source data for this study have been provided with available URLs

## Additional information

## Funding

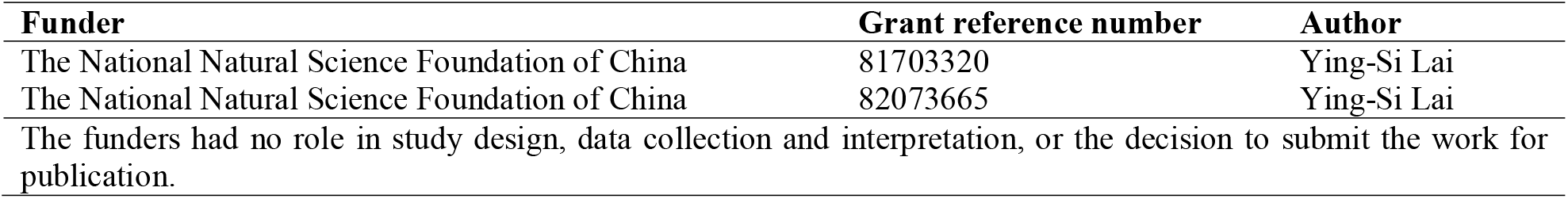

## Author contributions

HYX and YSL made substantial contributions to the study concept and design. HYX and YYZ made substantial contributions to data collection. TTZ and YSL made substantial contributions to data analysis, and interpretation. HYX, and YSL were in charge of the manuscript draft. JYC and YSL made substantial revisions to the manuscript.

## Acknowledgments

We are grateful to TTZ for providing very good suggestions for the data collection.

