## Appendix for "The spatial-temporal risk profiling of *Clonorchis sinensis* infection in South Korea: A geostatistical modeling study"

**Supplementary File 1: PRISMA 2020 for Checklist**

| Section and Topic | Item # | Checklist item | Location where item is reported |
|---|---|---|---|
| **TITLE** | | | |
| Title | 1 | Identify the report as a systematic review. | Title (page 1) |
| ABSTRACT | | | |
| Abstract | 2 | See the PRISMA 2020 for Abstracts checklist. | Supplementary File 2 |
| INTRODUCTION | | | |
| Rationale | 3 | Describe the rationale for the review in the context of existing knowledge. | Introduction (page 2-4) |
| Objectives | 4 | Provide an explicit statement of the objective(s) or question(s) the review addresses. | Introduction (page 4) |
| METHODS | | | |
| Eligibility criteria | 5 | Specify the inclusion and exclusion criteria for the review and how studies were grouped for the syntheses. | Method (“Disease data”: page 5), Supplementary Figure 3 |
| Information sources | 6 | Specify all databases, registers, websites, organisations, reference lists and other sources searched or consulted to identify studies. Specify the date when each source was last searched or consulted. | Method (“Disease data”: page 5), Supplementary Figure 3 |
| Search strategy | 7 | Present the full search strategies for all databases, registers and websites, including any filters and limits used. | Method (“Disease data”: page 4), Supplementary Figure 3 |
| Selection process | 8 | Specify the methods used to decide whether a study met the inclusion criteria of the review, including how many reviewers screened each record and each report retrieved, whether they worked independently, and if applicable, details of automation tools used in the process. | Method (“Disease data”: page 5), Supplementary |

| Section and Topic | Item # | Checklist item | Location where item is reported |
|---|---|---|---|
| | | | Figure 3 |
| Data collection process | 9 | Specify the methods used to collect data from reports, including how many reviewers collected data from each report, whether they worked independently, any processes for obtaining or confirming data from study investigators, and if applicable, details of automation tools used in the process. | Method ("Disease data": page 5), Supplementary Figure 3 |
| Data items | 10a | List and define all outcomes for which data were sought. Specify whether all results that were compatible with each outcome domain in each study were sought (e.g. for all measures, time points, analyses), and if not, the methods used to decide which results to collect. | Method ("Disease data": page 5), Supplementary Figure 3 |
| | 10b | List and define all other variables for which data were sought (e.g. participant and intervention characteristics, funding sources). Describe any assumptions made about any missing or unclear information. | Method ("Socioeconomic, environmental and demographic data": page 5-6), Supplementary Figure 4 |
| Study risk of bias assessment | 11 | Specify the methods used to assess risk of bias in the included studies, including details of the tool(s) used, how many reviewers assessed each study and whether they worked independently, and if applicable, details of automation tools used in the process. | Method ("Disease data": page 5), Supplementary File 5 |
| Effect measures | 12 | Specify for each outcome the effect measure(s) (e.g. risk ratio, mean difference) used in the synthesis or presentation of results. | Method ("Disease data": page 5) |
| Synthesis methods | 13a | Describe the processes used to decide which studies were eligible for each synthesis (e.g. tabulating the study intervention characteristics and comparing against the planned groups for each synthesis (item #5)). | Method ("Statistical analysis": page 6-7), Supplementary File 5 |
| | 13b | Describe any methods required to prepare the data for presentation or synthesis, such as handling of missing summary statistics, or data | Method |

| Section and Topic | Item # | Checklist item | Location where item is reported |
|---|---|---|---|
| | | conversions. | ("Statistical analysis": page 6-7), Supplementary File 5 |
| | 13c | Describe any methods used to tabulate or visually display results of individual studies and syntheses. | Method ("Statistical analysis": page 7) |
| | 13d | Describe any methods used to synthesize results and provide a rationale for the choice(s). If meta-analysis was performed, describe the model(s), method(s) to identify the presence and extent of statistical heterogeneity, and software package(s) used. | Method ("Statistical analysis": page 6-7), Supplementary File 5 |
| | 13e | Describe any methods used to explore possible causes of heterogeneity among study results (e.g. subgroup analysis, meta-regression). | Method ("Statistical analysis": page 6-7), Supplementary File 5 |
| | 13f | Describe any sensitivity analyses conducted to assess robustness of the synthesized results. | Method ("Statistical analysis": page 7), Supplementary File 7, Supplementary File 6, Supplementary File 7 |
| Reporting bias assessment | 14 | Describe any methods used to assess risk of bias due to missing results in a synthesis (arising from reporting biases). | Supplementary File 3 |
| Certainty assessment | 15 | Describe any methods used to assess certainty (or confidence) in the body of evidence for an outcome. | Method ("Statistical |

| Section and Topic | Item # | Checklist item | Location where item is reported |
|---|---|---|---|
| | | | analysis”: page 6-7), Supplementary File 5 |
| RESULTS | | | |
| Study selection | 16a | Describe the results of the search and selection process, from the number of records identified in the search to the number of studies included in the review, ideally using a flow diagram. | Figure 1 |
| | 16b | Cite studies that might appear to meet the inclusion criteria, but which were excluded, and explain why they were excluded. | Figure 1 |
| Study characteristics | 17 | Cite each included study and present its characteristics. | Results (“Data summaries”: page 7-8) |
| Risk of bias in studies | 18 | Present assessments of risk of bias for each included study. | Figure 3 |
| Results of individual studies | 19 | For all outcomes, present, for each study: (a) summary statistics for each group (where appropriate) and (b) an effect estimate and its precision (e.g. confidence/credible interval), ideally using structured tables or plots. | Table1, Figure 3 |
| Results of syntheses | 20a | For each synthesis, briefly summarise the characteristics and risk of bias among contributing studies. | Table1, Figure 3 |
| | 20b | Present results of all statistical syntheses conducted. If meta-analysis was done, present for each the summary estimate and its precision (e.g. confidence/credible interval) and measures of statistical heterogeneity. If comparing groups, describe the direction of the effect. | Table2 |
| | 20c | Present results of all investigations of possible causes of heterogeneity among study results. | Supplementary Table 3, Supplementary Table 4, Table1, Figure 3 |
| | 20d | Present results of all sensitivity analyses conducted to assess the robustness of the synthesized results. | Supplementary Table 3, Supplementary Table 4 |
| Reporting biases | 21 | Present assessments of risk of bias due to missing results (arising from reporting biases) for each synthesis assessed. | Supplementary Table 3, Supplementary Table 4 |

### PRISMA 2020 for Abstracts Checklist

| Section and Topic | Item # | Checklist item | Location where item is reported |
|---|---|---|---|
| Certainty of evidence | 22 | Present assessments of certainty (or confidence) in the body of evidence for each outcome assessed. | Results (Risk maps and estimates of number of people infected: page 14) Figure 5 |
| DISCUSSION | | | |
| Discussion | 23a | Provide a general interpretation of the results in the context of other evidence. | Discussion (page 18-21) |
| | 23b | Discuss any limitations of the evidence included in the review. | Discussion (page 20-21) |
| | 23c | Discuss any limitations of the review processes used. | Discussion (page 20-21) |
| | 23d | Discuss implications of the results for practice, policy, and future research. | Conclusion |
| OTHER INFORMATION | | | |
| Registration and protocol | 24a | Provide registration information for the review, including register name and registration number, or state that the review was not registered. | Method ("Disease data": page4) |
| | 24b | Indicate where the review protocol can be accessed, or state that a protocol was not prepared. | Supplementary Figure 3 |
| | 24c | Describe and explain any amendments to information provided at registration or in the protocol. | Supplementary Figure 3 |
| Support | 25 | Describe sources of financial or non-financial support for the review, and the role of the funders or sponsors in the review. | Funding (page23) |
| Competing interests | 26 | Declare any competing interests of review authors. | Competing interests (page23) |
| Availability of data, code and other materials | 27 | Report which of the following are publicly available and where they can be found: template data collection forms; data extracted from included studies; data used for all analyses; analytic code; any other materials used in the review. | Supplementary Table 1 |

*From:* Page MJ, McKenzie JE, Bossuyt PM, Boutron I, Hoffmann TC, Mulrow CD, et al. The PRISMA 2020 statement: an updated guideline for reporting systematic reviews. BMJ 2021;372:n71.

### PRISMA 2020 for Abstracts Checklist

doi: 10.1136/bmj.n71

For more information, visit: http://www.prisma-statement.org/

**Supplementary File 2: PRISMA 2020 for Abstracts Checklist**

| Section and Topic | Item # | Checklist item | Reported (Yes/No) |
|---|---|---|---|
| **TITLE** | | | |
| Title | 1 | Identify the report as a systematic review. | Yes |
| **BACKGROUND** | | | |
| Objectives | 2 | Provide an explicit statement of the main objective(s) or question(s) the review addresses. | Yes |
| **METHODS** | | | |
| Eligibility criteria | 3 | Specify the inclusion and exclusion criteria for the review. | Yes |
| Information sources | 4 | Specify the information sources (e.g. databases, registers) used to identify studies and the date when each was last searched. | Yes |
| Risk of bias | 5 | Specify the methods used to assess risk of bias in the included studies. | Yes |
| Synthesis of results | 6 | Specify the methods used to present and synthesise results. | Yes |
| **RESULTS** | | | |
| Included studies | 7 | Give the total number of included studies and participants and summarise relevant characteristics of studies. | Yes |
| Synthesis of results | 8 | Present results for main outcomes, preferably indicating the number of included studies and participants for each. If meta-analysis was done, report the summary estimate and confidence/credible interval. If comparing groups, indicate the direction of the effect (i.e. which group is favoured). | Yes |
| **DISCUSSION** | | | |
| Limitations of evidence | 9 | Provide a brief summary of the limitations of the evidence included in the review (e.g. study risk of bias, inconsistency and imprecision). | Yes |
| Interpretation | 10 | Provide a general interpretation of the results and important implications. | Yes |
| **OTHER** | | | |
| Funding | 11 | Specify the primary source of funding for the review. | Yes |
| Registration | 12 | Provide the register name and registration number. | Yes |

*From:* Page MJ, McKenzie JE, Bossuyt PM, Boutron I, Hoffmann TC, Mulrow CD, et al. The PRISMA 2020 statement: an updated guideline for reporting systematic reviews. BMJ 2021;372:n71.

doi: 10.1136/bmj.n71

For more information, visit: http://www.prisma-statement.org/

**Supplementary File 3: Quality assessment of eligible studies**

We did quality evaluation for each literature included in the final geostatistical modeling analysis, which is undertaken using an adapted nine-point quality assessment checklist (***Zhao et al., 2021***). The items of quality evaluation are as follows:

Q1: provide specific inclusion and exclusion criteria.

Q2: provide basic characteristics of the investigated population (e.g., gender, age, etc.).

Q3: provide prevalence rate of the survey.

Q4: provide number of positive patients and number of examined people of the survey.

Q5: provide diagnostic method used in the survey.

Q6: provide survey type.

Q7: provide time of the survey.

Q8: describe or discuss the possible bias of the survey or how confounders are controlled.

Q9: the literature comes from Science Citation Index Expanded database or Korea Citation Index.

1 point is assigned to the publication matching the item or 0 in contrary. The scores are summed up for all items and assigned to the publication as its quality score. The score for each literature is listed in Supplementary Table 1.

**Supplementary File 4: GATHER checklist of information that should be included in reports of global health estimates**

| # | Checklist item | Reported on page # |
|---|---|---|
| **Objectives and funding** | | |
| 1 | Define the indicator(s), populations (including age, sex, and geographic entities), and time period(s) for which estimates were made. | 5 |
| 2 | List the funding sources for the work. | 21 |
| **Data inputs** | | |
| For all data inputs from multiple sources that are synthesized as part of the study: | | |
| 3 | Describe how the data were identified and how the data were accessed. | 4-5 |
| 4 | Specify the inclusion and exclusion criteria. Identify all ad-hoc exclusions. | 5; Supplementary Figure 3 |
| 5 | Provide information about all included data sources and their main characteristics. For each data source used, report reference information or contact name/institution, population represented, data collection method, year(s) of data collection, sex and age range, diagnostic criteria or measurement method, and sample size, as relevant. | 4-6, Supplementary Table 1, Supplementary Figure 3 |
| 6 | Identify and describe any categories of input data that have potentially important biases (e.g., based on characteristics listed in item 5). | Table1; Figure 2, Figure 3, Supplementary Figure 7, Supplementary Figure 8, Supplementary Table 3, Supplementary Table 4 |
| For data inputs that contribute to the analysis but were not synthesized as part of the study: | | |
| 7 | Describe and give sources for any other data inputs. | Supplementary Table 1,Supplementary Table 5, |
| For all data inputs: | | |
| 8 | Provide all data inputs in a file format from which data can be efficiently extracted (e.g., a spreadsheet rather than a PDF), including all relevant meta-data listed in item 5. For any data inputs that cannot be shared because of ethical or legal reasons, such as third-party ownership, provide a contact name or the name of the institution that retains the right to the data. | Supplementary Table 1 |
| **Data analysis** | | |
| 9 | Provide a conceptual overview of the data analysis method. A diagram may be helpful. | 5-7 |
| 10 | Provide a detailed description of all steps of the analysis, including mathematical formulae. This description should cover, as relevant, data cleaning, data pre-processing, data adjustments and weighting of data sources, and mathematical or statistical model(s). | 5-7 |
| 11 | Describe how candidate models were evaluated and how the final model(s) were selected. | 7 |
| 12 | Provide the results of an evaluation of model performance, if done, as well as the results of any relevant sensitivity analysis. | 14; Supplementary Figure |

| | | |
|---|---|---|
| | | 7, Supplementary Figure 8, Supplementary Table 3, Supplementary Table 4 |
| 13 | Describe methods of calculating uncertainty of the estimates. State which sources of uncertainty were, and were not, accounted for in the uncertainty analysis. | 5-7 |
| 14 | State how analytic or statistical source code used to generate estimates can be accessed. | 5-7 |
| **Results and discussion** | | |
| 15 | Provide published estimates in a file format from which data can be efficiently extracted. | Table 1, Figure 3 |
| 16 | Report a quantitative measure of the uncertainty of the estimates (e.g., uncertainty intervals). | Figure 5 |
| 17 | Interpret results in light of existing evidence. If updating a previous set of estimates, describe the reasons for changes in estimates. | 18-20 |
| 18 | Discuss limitations of the estimates. Include a discussion of any modelling assumptions or data limitations that affect interpretation of the estimates. | 20-21 |

*From:* Stevens GA, Alkema L, Black RE, Boerma JT, Collins GS, Ezzati M, Grove JT, Hogan DR, Hogan MC, Horton R, Lawn JE, Marušić A, Mathers CD, Murray CJL, Rudan I, Salomon JA, Simpson PJ, Vos T, Welch V. 2016. Guidelines for Accurate and Transparent Health Estimates Reporting: the GATHER statement. *PLOS Medicine* **13**:e1002116. doi:10.1371/journal.pmed.1002056

**Supplementary File 5: Model fitting and variable selection**

A spatial-temporal model combined with covariates and an exchangeable non-spatial random effect was developed, which was defined as an autoregressive stochastic process with first order (AR1) model in the time domain and an SPDE (Stochastic Partial Differential Equation) model in the spatial dimension (***Cameletti et al., 2013; Krainski et al., 2019***). We assumed the infection risk remains unchanged in the same areas within 1-year period. Different observations from the same year and areas can be regarded as a randomized spatial-temporal process. In addition, we overlaid the survey area with a standard grid of $5 \times 5\ \mathrm{km}^2$ to generate 5946 pixels representing this area. survey locations and pixels within survey areas are assumed to share the same spatial-temporal process.

For survey data reporting both the number of examined and positive, we defined that $p_{it}, Y_{it}$ and $N_{it}$ are the number of examined individuals, the number of those positive and the probability of infection for location $i$ and in time period *t*. We assume that $Y_{it}$ followed a binominal distribution $Y_{it} \sim Bin(p_{it}, N_{it})$. For survey data only reporting observed prevalence, we assumed that the observed prevalence $ob_{it}$ arises from a beta distribution $ob_{it} \sim Be(p_{it}, \sigma_\beta^2)$. All covariates were modeled on the logit scale of $p_{it}$.

We defined that $i = 1, \ldots, n_A, n_{A+1}, \ldots. n_A + n_p$, where $i$ is the index of the survey location in point-level data, or the index of the survey area in areal level data, $n_A$ and $n_p$ refer to the total number of areas for areal level surveys and locations for point-level surveys, respectively). For areal level data, the Bayesian model is $Logit(p_{it}) = \beta_0 + \widetilde{x'_{it}}\beta + |A_i|^{-1} \int_{Ai} \boldsymbol{\omega}(s,t)\, dxdt + \varphi_i$, where $i = 1, \ldots, n_A$, $\beta_0$ and $\beta$ are the corresponding regression coefficients of the intercept and covariates (the vectors of covariate values $\widetilde{x'_{it}}$ for $i^{th}$ area in time period $t$ is denoted as $|A_i|^{-1} \int_{Ai} x(s,t)\, dxdt$), $|A_i|$=$\int_{Ai} 1ds$ means the size of the $i^{th}$ area), $\boldsymbol{\omega}(s,t)$ is the spatial-temporal random effects of pixels within the area, and $\varphi_i$ is the exchangeable non-spatial random effect following a zero-mean normal distribution $\varphi_i \sim N(0, \sigma_{nonsp}^2)$. For point-level data, the Bayesian model is somewhat different from above, that is $Logit(p_{it}) = \beta_0 + x'_{it}\beta + \boldsymbol{\omega}(s,t) + \varphi_i$. Here $i = n_{A+1}, \ldots. n_A + n_p$, $x'_{it}$ denotes the vectors of covariate values, and $\boldsymbol{\omega}(s_i, t)$ is the spatial-temporal random effects at the $i^{th}$ location and time period $t$. Assuming that the spatio-temporal random effect arises from a zero-mean Gaussian distribution, where $\boldsymbol{\omega} \sim GP(0, \boldsymbol{K}_{space} \otimes \boldsymbol{K}_{time})$. The term $\boldsymbol{K}_{space}$ is the spatial covariance matrix following a stationary Matérn covariance function $\sigma_\phi^2(\kappa\boldsymbol{D})^v K_v(\kappa\boldsymbol{D})/(\Gamma(v)2^{v-1})$, where $\sigma_\phi^2$ represents the spatial variance, $\kappa$ is a positive scaling parameter related to the spatial range $r = \sqrt{8v}/\kappa$, $r$ denotes the distance at which spatial correlation becomes negligible [<0.1]), $\boldsymbol{D}$ is Euclidean distance matrix, $K_v$ represents the modified Bessel function of the second kind defined by a smoothness parameter $v$ fixed to 1. $\boldsymbol{K}_{time}$ is the temporal covariance matrix, that is, $\boldsymbol{K}_{time} = \rho^{|t_u - t_o|}$,where $|\rho| < 1$, corresponding to AR1. In addition, we assumed the spatio-temporal random effect $\omega$ is mutually independent at each other in different locations ($i$ or $j$) and times ($t$ or $t'$), where $Cov(\omega_{it,}\omega_{jt'}) = \begin{cases} 0, & if\ t \neq t' \\ \sigma_\phi^2, & if\ t = t' \end{cases}$. The survey period of this study was from 1970 to 2017. We built the Gaussian Markov random field (GMRF) on regular temporal knots under the SPDE framework to decrease the computational burden, that is, $\omega = (\omega_t = 1970, \omega_t = 1977, \omega_t = 1985, \omega_t = 1993, \omega_t = 2001, \omega_t = 2009, \omega_t = 2017)$. The latent fields in other years are approximated by employing the B-spline basis function of degree two for the projection of $\omega$, that is $B_{i,1}(t) = \begin{cases} 1, & t_i \leq t < t_{i+1} \\ 0, & \text{otherwise} \end{cases}$ and $B_{i,m}(t) = \frac{t - t_i}{t_{i+m-1} - t_i} B_{i,m-1}(t) + \frac{t_{i+m} - t}{t_{i+m} - t_{i+1}} B_{i+1,m-1}(t)$, where the degree $m$ is equal to 2 (***Cameletti et al., 2013; Krainski et al., 2019***). The models were built under a Bayesian framework. Minimally informative priors were specified for regression coefficients, the precision parameters, temporal correlation coefficient, range parameter as follows: $\boldsymbol{\beta} \sim N(0, 10^5 \boldsymbol{I})$, $\log(1/\sigma_\phi^2) \sim \text{logGamma}(1, 0.01)$, $\log(1/\sigma_{nonsp}^2) \sim \text{logGamma}(1, 0.01)$, $\log((1+\rho)/(1-\rho)) \sim N(0, 0.15)$, $\log(\kappa) \sim N(\log(\sqrt{8}/d), 1)$, where $d$ is the median of distances between predicted grids.

#### Supplementary File 6: Model validation

5-fold out-of-sample cross-validation approach was conducted for model validation. we randomly partitioned all survey data into five subsets with similar sample size, of which four subsets were used as training data and the rest as validation data. The advanced Bayesian geostatistical model was fitted based on training data and the estimated prevalence was calculated on the locations of validation data. We calculated mean error ($ME = \frac{1}{N}\sum(es_{it} - ob_{it})$, where $N$ is the number of total data, $es_{it}$ is the estimated prevalence), mean absolute error ($\mathrm{MAE} = \frac{1}{N}\sum|es_{it} - ob_{it}|$), mean square error (($\mathrm{MSE} = \frac{1}{N}\sum(es_{it} - ob_{it})^2$)), the percentage of observations covered by 95% Bayesian credible intervals (BCIs) of posterior estimated prevalence**(*Brooker et al., 2001; Lai et al., 2015; Zhao et al., 2021*)**. The process was repeated five times, and we chose one different subset as the validation data each time, i.e., each subset should be the validation data once. In addition, the results were assessed under the receiver operating characteristic (ROC) curves. The results of five validations were merged, and a series of cut-off values from 0 to 1 were set, on the basis of which the observed and estimated prevalence was categorized as a binary number of 0 or 1. The percentage of surveys with both observed and estimated prevalence categorized to one was identified as the predictive accuracy with sensitivity, while the percentage of surveys with both observed and estimated prevalence categorized to zero was identified as specificity. We calculated the area under the receiver-operating characteristic (ROC) curve (AUC) to illustrate the prediction accuracy of our model by the AUC function in the package "DescTools" in R (version 4.0.4).

**Supplementary File 7: Sensitivity analysis for different diagnostic methods**

Considering current studies on the diagnostic abilities of different methods are controversial(***Cho et al., 1969; Hong et al., 2003; Uga et al., 2010***), we didn't take into account the effect of different diagnostic methods in the final model. To evaluate the effect of different diagnostic methods on our model, we conducted a sensitivity analysis. Cho et al's atudy showed that the prevalence obtained by Formalin-ether sedimentation technique (FE) was 1.32 times higher than Kato-Katz **(*Cho et al., 1969*)**. We assumed that the original number of examined of each survey is unchanged. to avoid a situation that the number of positive greater than the examined, we divided the prevalence obtained by FE by 1.32 as the adjusted prevalence. For survey data reporting both the number of examined and positive, the adjusted number of positive is equal to the number of examined multiplied by the adjusted prevalence. We assume that the adjusted number of those positive $\acute{Y}_{it}$ followed a binominal distribution $\acute{Y}_{it} \sim Bin(\acute{p}_{it}, N_{it})$, where $\acute{p}_{it}$ is the adjusted prevalence. For data only reporting observed prevalence, we assumed that the observed prevalence $ob_{it}$ arises from a beta distribution $ob_{it} \sim Be(\acute{p}_{it}, \sigma_{\beta}^{2})$. In this way, the number of examined and positive included in the model were obtained based on Kato-Katz. In addition, to estimate the maximum possible prevalence in South Korea, we considered 1.32 times of the estimated prevalence as the adjusted estimated prevalence for the risk maps. Finally, adjusted and unadjusted results were compared.

**Supplementary File 8: Sensitivity analysis for surveys reported prevalence in intervals**

In the final model, three publications including seven surveys reported observed prevalence of *C. sinensis* infection in intervals for 186 area level data and 36 point-referenced disease across South Korea (***Korea Centers for Disease Control and Prevention and Korea National Institute of Health. 2013; Cho et al., 2016; Ju et al., 2005***). To make full use of all eligible data, we used the median values of the intervals as observed prevalence for our final model. Sensitivity analysis was done by using the lower and the upper limits of the intervals in the modeling analysis and results were compared.

**Supplementary Table 1: Result of quality assessment for eligible literatures**

| Numbers of the publication | Literature (author & year of publication) | Q1 | Q2 | Q3 | Q4 | Q5 | Q6 | Q7 | Q8 | Q9 | Total score |
|---|---|---|---|---|---|---|---|---|---|---|---|
| 1 | **(*Kim et al., 1971*)** | 1 | 1 | 1 | 1 | 1 | 1 | 1 | 0 | 1 | 8 |
| 2 | **(*The Ministry of Health and Social Affairs and Korean Association of parasite Eradication. 1971*)** | 0 | 0 | 1 | 1 | 1 | 0 | 0 | 0 | 0 | 3 |
| 3 | **(*Kang, 1972*)** | 1 | 1 | 1 | 1 | 1 | 1 | 1 | 0 | 0 | 7 |
| 4 | **(*Choi et al., 1973*)** | 1 | 1 | 1 | 1 | 1 | 1 | 1 | 0 | 1 | 8 |
| 5 | **(*Joo & Choi, 1974*)** | 1 | 1 | 1 | 1 | 1 | 1 | 1 | 0 | 1 | 8 |
| 6 | **(*Lee & Loh, 1974*)** | 1 | 1 | 1 | 1 | 1 | 1 | 1 | 0 | 0 | 7 |
| 7 | **(*Kim & Yu, 1975*)** | 1 | 1 | 1 | 1 | 1 | 1 | 1 | 1 | 0 | 8 |
| 8 | **(*Lee et al., 1975*)** | 1 | 1 | 1 | 1 | 1 | 1 | 1 | 0 | 1 | 8 |
| 9 | **(*Choi et al., 1976*)** | 1 | 1 | 1 | 1 | 1 | 1 | 1 | 0 | 1 | 8 |
| 10 | **(*Soh et al., 1976*)** | 1 | 1 | 1 | 1 | 1 | 1 | 1 | 0 | 0 | 7 |
| 11 | **(*The Ministry of Health and Social Affairs and The Korean Association of parasite Eradication. 1976*)** | 1 | 1 | 1 | 1 | 1 | 1 | 1 | 1 | 0 | 8 |
| 12 | **(*Chai et al., 1977*)** | 0 | 1 | 1 | 1 | 1 | 1 | 1 | 0 | 0 | 6 |
| 13 | **(*Shin, 1977*)** | 1 | 1 | 1 | 1 | 1 | 1 | 1 | 1 | 0 | 8 |
| 14 | **(*Soh & Ahn, 1978*)** | 1 | 1 | 1 | 1 | 1 | 1 | 1 | 1 | 1 | 9 |
| 15 | **(*Choi, 1978*)** | 1 | 1 | 1 | 1 | 1 | 1 | 1 | 0 | 1 | 8 |
| 16 | **(*Kang, 1978*)** | 1 | 1 | 1 | 1 | 1 | 1 | 1 | 0 | 1 | 8 |
| 17 | **(*Rim et al., 1978*)** | 1 | 1 | 1 | 1 | 1 | 1 | 1 | 0 | 0 | 7 |
| 18 | **(*Moon et al., 1979*)** | 1 | 1 | 1 | 1 | 1 | 1 | 1 | 0 | 1 | 8 |
| 19 | **(*Ban, 1979*)** | 1 | 1 | 1 | 1 | 1 | 1 | 1 | 0 | 0 | 7 |
| 20 | **(*Kim, 1980*)** | 1 | 1 | 1 | 1 | 1 | 1 | 1 | 0 | 1 | 8 |
| 21 | **(*Chang et al., 1980*)** | 1 | 1 | 1 | 1 | 1 | 1 | 1 | 0 | 0 | 7 |
| 22 | **(*Joo, 1980*)** | 1 | 1 | 1 | 1 | 1 | 1 | 1 | 0 | 0 | 7 |
| 23 | **(*Seo et al., 1981*)** | 1 | 1 | 1 | 1 | 1 | 1 | 1 | 0 | 0 | 7 |
| 24 | **(*Moon et al., 1981*)** | 1 | 1 | 1 | 1 | 1 | 1 | 1 | 0 | 0 | 7 |
| 25 | **(*Kim & Guh, 1981*)** | 1 | 1 | 1 | 1 | 1 | 1 | 1 | 0 | 0 | 7 |
| 26 | **(*Ryu et al., 1981*)** | 1 | 1 | 1 | 1 | 1 | 1 | 1 | 0 | 0 | 7 |
| 27 | **(*Song et al., 1981*)** | 1 | 1 | 1 | 1 | 1 | 1 | 1 | 1 | 0 | 8 |

| Numbers of the publication | Literature (author & year of publication) | Q1 | Q2 | Q3 | Q4 | Q5 | Q6 | Q7 | Q8 | Q9 | Total score |
|---|---|---|---|---|---|---|---|---|---|---|---|
| 28 | **(*Choi & Kim, 1981*)** | 1 | 1 | 1 | 1 | 1 | 1 | 1 | 0 | 0 | 7 |
| 29 | **(*The Ministry of Health and Social Affairs and The Korea Association of Parasite Eradication. 1982*)** | 1 | 1 | 1 | 1 | 1 | 1 | 1 | 1 | 0 | 8 |
| 30 | **(*Joo, 1981*)** | 1 | 1 | 1 | 1 | 1 | 1 | 1 | 0 | 0 | 7 |
| 31 | **(*Song, 1982*)** | 1 | 1 | 1 | 1 | 1 | 1 | 1 | 0 | 1 | 8 |
| 32 | **(*Joo et al., 1982*)** | 1 | 1 | 1 | 1 | 1 | 1 | 1 | 1 | 0 | 8 |
| 33 | **(*Rim et al., 1982*)** | 1 | 1 | 1 | 1 | 1 | 1 | 1 | 0 | 0 | 7 |
| 34 | **(*Cho et al., 1982*)** | 1 | 1 | 1 | 1 | 1 | 1 | 1 | 0 | 1 | 8 |
| 35 | **(*Moon et al., 1982*)** | 1 | 1 | 1 | 1 | 1 | 1 | 1 | 0 | 0 | 7 |
| 36 | **(*Park, 1982*)** | 1 | 1 | 1 | 1 | 1 | 1 | 1 | 0 | 0 | 7 |
| 37 | **(*Song et al., 1983*)** | 1 | 1 | 1 | 1 | 1 | 1 | 1 | 0 | 0 | 7 |
| 38 | **(*Kim, 1983*)** | 1 | 1 | 1 | 1 | 1 | 1 | 1 | 0 | 0 | 7 |
| 39 | **(*Rhee et al., 1983*)** | 1 | 1 | 1 | 1 | 1 | 1 | 1 | 1 | 1 | 9 |
| 40 | **(*Joo, 1984*)** | 1 | 1 | 1 | 1 | 1 | 1 | 1 | 1 | 1 | 9 |
| 41 | **(*Choi et al., 1984*)** | 1 | 1 | 1 | 1 | 1 | 1 | 1 | 0 | 1 | 8 |
| 42 | **(*Kim et al., 1984*)** | 1 | 1 | 1 | 1 | 1 | 1 | 0 | 0 | 0 | 6 |
| 43 | **(*Ahn, 1984*)** | 1 | 1 | 1 | 1 | 1 | 1 | 1 | 0 | 1 | 8 |
| 44 | **(*Kim, 1984*)** | 1 | 1 | 1 | 1 | 1 | 1 | 1 | 0 | 0 | 7 |
| 45 | **(*Soh et al., 1985*)** | 1 | 0 | 1 | 1 | 1 | 1 | 1 | 0 | 0 | 6 |
| 46 | **(*Rim, 1985*)** | 1 | 1 | 1 | 1 | 1 | 1 | 1 | 0 | 0 | 7 |
| 47 | **(*Lee et al., 1985*)** | 1 | 1 | 1 | 1 | 1 | 1 | 0 | 0 | 0 | 6 |
| 48 | **(*Chung et al., 1986*)** | 1 | 1 | 1 | 1 | 1 | 1 | 1 | 1 | 0 | 8 |
| 49 | **(*Jeong, 1986*)** | 1 | 1 | 1 | 1 | 1 | 1 | 1 | 0 | 0 | 7 |
| 50 | **(*Joo & Baik, 1986*)** | 0 | 0 | 0 | 1 | 1 | 0 | 0 | 0 | 0 | 2 |
| 51 | **(*The Ministry of Health and Welfare and Korea Association of Health Promotion. 1997a*)** | 1 | 1 | 1 | 1 | 1 | 1 | 1 | 0 | 0 | 7 |
| 52 | **(*Ha & Joo, 1987*)** | 1 | 1 | 1 | 1 | 1 | 1 | 1 | 0 | 1 | 8 |
| 53 | **(*Joo et al., 1987*)** | 1 | 1 | 1 | 1 | 1 | 1 | 1 | 0 | 0 | 7 |
| 54 | **(*Joo et al., 1988*)** | 1 | 1 | 1 | 1 | 1 | 1 | 1 | 0 | 1 | 8 |
| 55 | **(*Hong et al., 1990*)** | 1 | 1 | 1 | 1 | 1 | 1 | 1 | 0 | 1 | 8 |
| 56 | **(*Chung et al., 1991*)** | 1 | 1 | 1 | 1 | 1 | 1 | 1 | 0 | 1 | 8 |
| 57 | **(*Yun, 1991*)** | 1 | 1 | 1 | 1 | 1 | 1 | 1 | 0 | 0 | 7 |

| Numbers of the publication | Literature (author & year of publication) | Q1 | Q2 | Q3 | Q4 | Q5 | Q6 | Q7 | Q8 | Q9 | Total score |
|---|---|---|---|---|---|---|---|---|---|---|---|
| 58 | (*The Ministry of Health and Social Affairs and The Korea Association of Health. 1993*) | 0 | 0 | 1 | 1 | 1 | 0 | 0 | 0 | 0 | 3 |
| 59 | (*Kim, 1992*) | 1 | 1 | 1 | 1 | 1 | 1 | 1 | 0 | 0 | 7 |
| 60 | (*Lee et al., 1993*) | 1 | 1 | 1 | 1 | 1 | 1 | 1 | 0 | 1 | 8 |
| 61 | (*Yu et al., 1994*) | 1 | 1 | 1 | 1 | 1 | 1 | 1 | 0 | 1 | 8 |
| 62 | (*Hong et al., 1994*) | 1 | 0 | 1 | 1 | 1 | 1 | 1 | 0 | 1 | 7 |
| 63 | (*Kim et al., 1994*) | 1 | 1 | 1 | 1 | 1 | 1 | 1 | 0 | 1 | 8 |
| 64 | (*Son et al., 1994*) | 1 | 0 | 1 | 1 | 1 | 1 | 1 | 0 | 1 | 7 |
| 65 | (*Choi & Joo, 1994*) | 1 | 1 | 1 | 1 | 1 | 1 | 1 | 0 | 1 | 8 |
| 66 | (*Lee et al., 1994a*) | 1 | 1 | 1 | 1 | 1 | 1 | 1 | 0 | 0 | 7 |
| 67 | (*Lee et al., 1994b*) | 1 | 1 | 1 | 1 | 1 | 1 | 1 | 0 | 0 | 7 |
| 68 | (*Lee et al., 1995*) | 1 | 1 | 1 | 1 | 1 | 1 | 1 | 0 | 0 | 7 |
| 69 | (*Kang, 1997*) | 1 | 1 | 1 | 1 | 1 | 1 | 1 | 0 | 1 | 8 |
| 70 | (*Hwang et al., 1997*) | 1 | 1 | 1 | 1 | 1 | 1 | 1 | 0 | 0 | 7 |
| 71 | (*The Ministry of Health and Welfare and Korea Association of Health Promotion. 1997b*) | 1 | 1 | 1 | 1 | 1 | 1 | 1 | 1 | 0 | 8 |
| 72 | (*Chai et al., 1998*) | 1 | 1 | 1 | 1 | 1 | 1 | 1 | 0 | 1 | 8 |
| 73 | (*Hong et al., 1998*) | 1 | 1 | 1 | 1 | 1 | 1 | 1 | 0 | 1 | 8 |
| 74 | (*Han et al., 1998*) | 1 | 1 | 1 | 1 | 1 | 1 | 1 | 0 | 1 | 8 |
| 75 | (*Shin et al., 1999*) | 1 | 1 | 1 | 1 | 1 | 1 | 1 | 1 | 1 | 9 |
| 76 | (*Kim et al., 1999*) | 1 | 1 | 1 | 1 | 1 | 1 | 1 | 0 | 0 | 7 |
| 77 | (*Park et al., 2000*) | 1 | 1 | 1 | 1 | 1 | 1 | 1 | 0 | 0 | 7 |
| 78 | (*Chai et al., 2000*) | 1 | 1 | 1 | 1 | 1 | 1 | 1 | 0 | 1 | 8 |
| 79 | (*Kim et al., 2001*) | 1 | 1 | 1 | 1 | 1 | 1 | 1 | 0 | 1 | 8 |
| 80 | (*Park et al., 2001*) | 0 | 1 | 1 | 1 | 1 | 1 | 1 | 0 | 1 | 7 |
| 81 | (*Sohn et al., 2001*) | 1 | 1 | 1 | 1 | 1 | 1 | 1 | 1 | 0 | 8 |
| 82 | (*B. J. Kim et al., 2002*) | 1 | 1 | 1 | 1 | 1 | 1 | 1 | 0 | 1 | 8 |
| 83 | (*Lee et al., 2002*) | 1 | 1 | 1 | 1 | 1 | 1 | 1 | 0 | 1 | 8 |
| 84 | (*S. I. Kim et al., 2002*) | 1 | 1 | 1 | 1 | 1 | 1 | 1 | 0 | 0 | 7 |
| 85 | (*Min et al., 2002*) | 1 | 1 | 1 | 1 | 1 | 1 | 1 | 0 | 1 | 8 |
| 86 | (*Joo, 2003*) | 1 | 1 | 1 | 1 | 1 | 1 | 1 | 0 | 1 | 8 |
| 87 | (*Kim & Yun, 2004*) | 1 | 1 | 1 | 1 | 1 | 1 | 1 | 1 | 1 | 9 |
| 88 | (*Korea Association of* | 1 | 1 | 1 | 1 | 1 | 1 | 1 | 1 | 0 | 8 |

| Numbers of the publication | Literature (author & year of publication) | Q1 | Q2 | Q3 | Q4 | Q5 | Q6 | Q7 | Q8 | Q9 | Total score |
|---|---|---|---|---|---|---|---|---|---|---|---|
| | ***Health Promotion. 2004*)** | | | | | | | | | | |
| 89 | **(*Ju et al., 2005*)** | 1 | 1 | 1 | 1 | 0 | 1 | 1 | 1 | 1 | 8 |
| 90 | **(*Guk et al., 2006*)** | 1 | 1 | 1 | 1 | 1 | 1 | 1 | 0 | 1 | 8 |
| 91 | **(*Lim et al., 2006*)** | 1 | 1 | 1 | 1 | 1 | 1 | 1 | 0 | 1 | 8 |
| 92 | **(*Park, Guk, et al., 2007*)** | 1 | 1 | 1 | 1 | 1 | 1 | 1 | 0 | 1 | 8 |
| 93 | **(*Park, Kim, et al., 2007*)** | 1 | 1 | 1 | 1 | 1 | 1 | 1 | 0 | 1 | 8 |
| 94 | **(*Shen et al., 2007*)** | 1 | 1 | 1 | 1 | 1 | 1 | 1 | 1 | 1 | 9 |
| 95 | **(*Kim, 2007*)** | 1 | 1 | 1 | 1 | 1 | 1 | 1 | 0 | 1 | 8 |
| 96 | **(*Park, 2007*)** | 1 | 1 | 1 | 1 | 1 | 1 | 1 | 1 | 1 | 9 |
| 97 | **(*Lee, 2008*)** | 1 | 1 | 1 | 1 | 1 | 1 | 1 | 1 | 0 | 8 |
| 98 | **(*Park, 2008*)** | 1 | 1 | 1 | 1 | 1 | 1 | 1 | 1 | 0 | 8 |
| 99 | **(*Cho et al., 2008*)** | 1 | 1 | 1 | 1 | 1 | 1 | 1 | 0 | 1 | 8 |
| 100 | **(*Lee et al., 2008*)** | 1 | 1 | 1 | 1 | 1 | 1 | 1 | 0 | 1 | 8 |
| 101 | **(*Cho et al., 2010*)** | 1 | 1 | 1 | 1 | 1 | 1 | 1 | 1 | 1 | 9 |
| 102 | **(*Shin et al., 2010*)** | 0 | 1 | 1 | 1 | 1 | 1 | 1 | 1 | 1 | 8 |
| 103 | **(*The Ministry of Health and Social Affairs. 2010*)** | 1 | 1 | 1 | 1 | 1 | 1 | 1 | 1 | 0 | 8 |
| 104 | **(*Choi et al., 2011*)** | 1 | 1 | 1 | 1 | 1 | 1 | 1 | 0 | 1 | 8 |
| 105 | **(*Korea Centers for Disease Control and Prevention and Korea National Institute of Health. 2013*)** | 1 | 1 | 1 | 0 | 0 | 1 | 1 | 1 | 0 | 6 |
| 106 | **(*June et al., 2013*)** | 1 | 1 | 1 | 1 | 1 | 1 | 1 | 0 | 1 | 8 |
| 107 | **(*Park et al., 2014*)** | 1 | 1 | 1 | 1 | 1 | 1 | 1 | 1 | 1 | 9 |
| 108 | **(*Oh et al., Blackwell Publishing Ltd. 2014*)** | 1 | 1 | 1 | 1 | 1 | 1 | 1 | 1 | 1 | 9 |
| 109 | **(*Cho et al., 2016*)** | 0 | 1 | 1 | 0 | 1 | 1 | 1 | 0 | 0 | 5 |
| 110 | **(*Jeong et al., 2017*)** | 1 | 1 | 1 | 1 | 1 | 1 | 1 | 1 | 1 | 9 |
| 111 | **(*Shin et al., 2017*)** | 1 | 1 | 1 | 1 | 1 | 1 | 1 | 0 | 0 | 7 |
| 112 | **(*Bahk et al., 2018*)** | 1 | 1 | 1 | 1 | 1 | 1 | 1 | 0 | 1 | 8 |

**Reference**

The Ministry of Health and Social Affairs, Korean Association of parasite Eradication. *Status of the first Korean intestinal parasite infection*. The Ministry of Health and Social Affairs, Korean Association of parasite Eradication; 1971.

The Ministry of Health and Social Affairs, The Korean Association of parasite Eradication. *Prevalence of Intestinal Parasitic Infections in Korea-The 2nd Report*. The Ministry of Health and Social Affairs, The Korean Association of parasite Eradication; 1976.

The Ministry of Health and Social Affairs, The Korea Association of Parasite Eradication. *Prevalence of Intestinal Parasitic Infections in Korea-The Third Report*. The Ministry of Health and Social Affairs, The Korea Association of Parasite Eradication; 1982.

The Ministry of Health and Social Affairs, The Korea Association of Health. *Prevalence of Intestinal Parasitic Infections in Korea-The Fifith Report*. The Ministry of Health and Social Affairs, The Korea Association of Health; 1993.

The Ministry of Health and Welfare, Korea Association of Health Promotion. *Prevalence of intestinal parasitic infections in Korea-The Sixth Report.* The Ministry of Health and Welfare, Korea Association of Health Promotion; 1997a.

The Ministry of Health and Welfare, Korea Association of Health Promotion. *Prevalence of Intestinal Parasitic Infections in Korea-The Sixth Report*. The Ministry of Health and Welfare, Korea Association of Health Promotion; 1997b.

Korea Association of Health Promotion. *Prevalence of Intestinal Parasitic Infections in Korea-The 7th Report*. Korea Association of Health Promotion; 2004.

The Ministry of Health and Social Affairs. *Evaluation for Effects and Utilization of Clonorchiasis Prevention Program among High Risk Population at 5 Big Rivers in Korea*. The Ministry of Health and Social Affairs; 2010.

Korea Centers for Disease Control and Prevention, Korea National Institute of Health. *National survey of the prevalence of Intestinal Parasitic Infections in Korea, the 8th Report*. Korea Centers for Disease Control and Prevention, Korea National Institute of Health; 2013.

Ahn YK. Epidemiological studies on Metagonimus yokogawai infection in Samcheok-gun, Kangwon-do, Korea. *the Korean Journal of Parasitology.* 1984*, 22*: 161-170. doi:10.3347/kjp.1984.22.2.161.

Bahk YY, Park YK, Na BK, Sohn WM, Hong SJ, Chai JY, & Kim TS. Survey on Intestinal Helminthic Infection Status of Students in Two Counties, Hadong-gun and Goseong-gun, Korea. *the Korean Journal of Parasitology.* 2018*, 56*: 335-339. doi:10.3347/kjp.2018.56.4.335.

Ban JC. *Epidemiological study of Metagonimus Yokogawai at the area of Tamjin River in Korea*. 1979; Gwangju. http://www.riss.kr/link?id=T6459364

Chai JY, Cho SY, & Seo BS. Study on Metagonimus yokogawai(Katsurada, 1912) in Korea: IV. An epidemiological investigation along Tamjin River basin, South Cholla Do, Korea. *the Korean*

*Journal of Parasitology.* 1977*, 15*: 115-120. doi:10.3347/kjp.1977.15.2.115.

Chai JY, Han ET, Park YK, Guk SM, Kim JL, & Lee SH. High endemicity of Metagonimus yokogawai infection among residents of Samchok-shi, Kangwon-do. *the Korean Journal of Parasitology.* 2000*, 38*: 33-36. doi:10.3347/kjp.2000.38.1.33.

Chai JY, Song TE, Han ET, Guk SM, Park YK, Choi MH, & Lee SH. Two endemic foci of heterophyids and other intestinal fluke infections in southern and western coastal areas in Korea. *the Korean Journal of Parasitology.* 1998*, 36*: 155-161. doi:10.3347/kjp.1998.36.3.155.

Chang DY, Kim SH, Aha SW, Kim GS, & Baik KH. A Study on the Epidemiology of Clonorchiasis in Gum River Basin and Chemotherapeutic Effects of Embay 8440 ( Praziquantel ) against Clonorehiasis. *The Korean Journal of Medicine.* 1980*, 23*: 995-1002.

Cho KJ, Yun OS, Choo SK, & Shin HS. An Epidemiological study on clonorcohiasis at a Gum-River Basin. *Journal of Environmental Health Sciences.* 1982*, 8*: 61-66.

Cho SH, Cho PY, Lee DM, Kim TS, Kim IS, Hwang EJ, Na BK, & Sohn WM. Epidemiological Survey on the Infection of Intestinal Flukes in Residents of Muan-gun, Jeollanam-do, the Republic of Korea. *the Korean Journal of Parasitology.* 2010*, 48*: 133-138. doi:10.3347/kjp.2010.48.2.133.

Cho SH, Lee KY, Lee BC, Cho PY, Cheun HI, Hong ST, Sohn WM, & Kim TS. Prevalence of clonorchiasis in southern endemic areas of Korea in 2006. *the Korean Journal of Parasitology.* 2008*, 46*: 133-137. doi:10.3347/kjp.2008.46.3.133.

Cho SH, Shin HE, Lee SE, & Park MY. Survey on the Prevalence of Intestinal Parasitic Infections in Korea, 2014. *Public Health Weekly Report.* 2016*, 9*: 118-124.

Choi DW. Prevalence of Clonorchis sinensis in vicinity of Seongju, Kyungpook Province, Korea. *the Korean Journal of Parasitology.* 1978*, 16*: 140-147. doi:10.3347/kjp.1978.16.2.140.

Choi DW, Ahn DH, Choy CH, & Kim SS. Clonorchis Sinensis In Kyungpook Province, Korea: 3. Changing Pattern Of Clonorchis Sinensis Infection Among Inhabitants. *the Korean Journal of Parasitology.* 1976*, 14*: 117-122. doi:10.3347/kjp.1976.14.2.117.

Choi DW, Joo CY, Park SD, & Kim JW. Changing Pattern Of Clonorchis Sinensis Infection Among School Children In The Gumho Basin, Kyungpook Province, Korea. *the Korean Journal of Parasitology.* 1973*, 11*: 26-32. doi:10.3347/kjp.1973.11.1.26.

Choi DW, & Kim JH. Intestinal Helminth Survey of School Children near the River Taechong. *The Kyungpook University Medical Journal.* 1981*, 22*: 179-184.

Choi KJ, & Joo CY. Recent patterns of Entameba histolytica and other intestinal parasite infections in Taegu, Korea. *The Kyungpook University Medical Journal.* 1994*, 13*: 151-162.

Choi MH, Chang YS, Lim MK, Bae YM, Hong ST, Oh JK, Yun EH, Bae MJ, Kwon HS, Lee SM, Park HW, Min KU, Kim YY, & Cho SH. Clonorchis sinensis infection is positively associated with atopy in endemic area. *Clinical and Experimental Allergy.* 2011*, 41*: 697-705. doi:10.1111/j.1365-2222.2011.03746.x.

Choi SC, Moon JG, & Chung YH. Epidemiological Investigation on Metagonimus yokogawai and Clonorchis sinensis Infections along Tamjin River Basin. *The Medical Journal of Chosun University.* 1984*, 9*: 175-186.

Chung DI, Kim YI, Lee KR, & Choi DW. Epidemiological studies of digenetic trematodes in Yongyang County, Kyungpook Province. *the Korean Journal of Parasitology.* 1991*, 29*: 325-338. doi:10.3347/kjp.1991.29.4.325.

Chung MS, Lee JS, Rim HJ, Yum YT, Cha CW, & Koo BH. Survey on the Status of C. sinensis Infection in Rural Ingabitants(Yeoju Eup, Kyunggi Do). *Korean Journal of Rural Medicine.* 1986*, 11*: 3-11.

Guk SM, Park JH, Shin EH, Kim JL, Lin A, & Chai JY. Prevalence of Gymnophalloides seoi infection in coastal villages of Haenam-gun and Yeongam-gun, Republic of Korea. *the Korean Journal of Parasitology.* 2006*, 44*: 1-5. doi:10.3347/kjp.2006.44.1.1.

Ha JH, & Joo CY. Epidemiological studies of Entameba histolytica and other intetinal Parasites in Ulchin county, Kyungpook Province, Korea. *Keimyung Medical Journal.* 1987*, 6*: 26-33.

Han KH, Park NJ, & Eom KS. An Epidemiological Survey of Helminthic Infections Among Inhabitants in Youngdong-Gun, Chungbuk. *Chungbuk Medical Journal.* 1998*, 8*: 21-27.

Hong SJ, Lee YH, Chung MH, Lee DH, & Woo HC. Egg positive rates of Clonorchis sinensis and intestinal helminths among residents in Kagye-ri, Saengbiryang-myon, Sanchong-gun, Kyongsangnam-do. *the Korean Journal of Parasitology.* 1994*, 32*: 271-273. doi:10.3347/kjp.1994.32.4.271.

Hong SJ, Woo HC, Han JH, & Seong YK. Intestinal parasite infections among inhabitants in two islands of Tongyeong-gun, Kyeongsangnam-do. *the Korean Journal of Parasitology.* 1990*, 28*: 63-67. doi:10.3347/kjp.1990.28.1.63.

Hong ST, Yoon K, Lee M, Seo M, Choi MH, Sim JS, Choi BI, Yun CK, & Lee SH. Control of clonorchiasis by repeated praziquantel treatment and low diagnostic efficacy of sonography. *the Korean Journal of Parasitology.* 1998*, 36*: 249-254. doi:10.3347/kjp.1998.36.4.249.

Hwang MH, Kim SI, Park J, Ryu SY, Lee CG, Ahn HO, Kim YO, & Kim KS. The Prevalence of Clonorchis sinensis and Its Related Factors at Goksung Area in the Basin of Sumjin River. *Korean Journal of Rural Medicine.* 1997*, 22*: 239-252.

Jeong JY, Lee JY, Chung BS, Choi Y, Alley AB, & Kim HJ. A new method for estimating the prevalence of clonorchiasis in Korea A proposal to replace arbitrary riverside sampling. *Medicine (Baltimore).* 2017*, 96*: e6536. doi:10.1097/md.0000000000006536.

Jeong S, W. A Survey on the Infection of Clonorchiasis in the Rural Area. *Journal of the Korean Institute of Health.* 1986*, 9*: 181-192.

Joo CY. Recent pattern of intestinal helminth infections among residents in Ulju county, Kyungnam Province, Korea. *The New medical journal.* 1981*, 24*: 83-92.

Joo CY. Recent patterns of intestinal helminth infections among the residents in Taegu City, Korea. *the Korean Journal of Parasitology.* 1984*, 22*: 109-115. doi:10.3347/kjp.1984.22.1.109.

Joo CY. Epidemiological Studies of Clonorchis sinensis in Vicinity of Hapcheon Dam, Gyungnam Province, Korea. *Keimyung Medical Journal.* 2003*, 22*: 195-206.

Joo CY, & Baik DH. Prevalence of intestinal helminth infections in Kyungpook Province, Korea. *The New medical journal.* 1986*, 29*: 93-102.

Joo CY, Baik DH, Kang CM, Kwon TC, & Kim SJ. Prevalence of Entameba histolytica in school children of Taegu city, Korea. *Keimyung Medical Journal.* 1988*, 7*: 219-226.

Joo CY, & Choi DW. Newly found endemic foci of Clonorchis sinensis in Kyungpook Province, Korea. *the Korean Journal of Parasitology.* 1974*, 12*: 111-118. doi:10.3347/kjp.1974.12.2.111.

Joo JY. Epidemiological survey of Clonorchis sinensis among residents in Chunseong County, Kangweon Province, Korea. *Modern Medicine.* 1980*, 23*: 129-135.

Joo KH, Choi DL, & Rim HJ. Epidemiological Survey on Clonorchis sinensis in Yeoju Gun Gyeong-gi Do. *Korean Journal of Rural Medicine.* 1982*, 7*: 43-49.

Joo KH, Chu PB, Rim HJ, & Lee JS. Studies on the Epidemiological Change of Clonorchiasis After Mass Chemotherapy in Highly Endemic Areas. *Korean Journal of Rural Medicine.* 1987*, 12*: 80-93.

Ju YH, Oh JK, Kong HJ, Sohn WM, Kim JI, Jung KY, Kim YG, & Shin HR. Epidemiologic study of Clonorchis sinensis infestation in a rural area of Kyongsangnam-do, South Korea. *Journal of Preventive Medicine & Public Health* 2005*, 38*: 425-430.

June KJ, Cho SH, Lee WJ, Kim C, & Park KS. Prevalence and Risk Factors of Clonorchiasis among the Populations Served by Primary Healthcare Posts along Five Major Rivers in South Korea. *Osong Public Health and Research Perspectives.* 2013*, 4*: 21-26. doi:10.1016/j.phrp.2012.12.002.

Kang SB. *Changing Patterns of Clonorchis Sinensis Infections in Yeongcheon, Kyungpook Provincek, Korea*. 1997; Daegu.

Kang SY. An Epidemiological Analysis of the Clonorchiasis in an Area of North Choong Chong Do(=province). *Korean Journal of Public Health.* 1972*, 9*: 105-112.

Kang YS. (A) Study on the Status of Intestinal Parasites Infection in the Pediatric Age Group. *The Medical Journal of Chosun University.* 1978;43-62.

Kim BJ, Ock MS, Kim IS, & Yeo UB. Infection status of Clonorchis sinensis in residents of Hamyang-gun, Gyeongsangnam-do, Korea. *the Korean Journal of Parasitology.* 2002*, 40*: 191-193. doi:10.3347/kjp.2002.40.4.191.

Kim BJ, Yeon JW, & Ock MS. Infection rates of Enterobius vermicularis and Clonorchis sinensis of primary school children in Hamyang-gun, Gyeongsangnam-do (province), Korea. *the Korean Journal of Parasitology.* 2001*, 39*: 323-325. doi:10.3347/kjp.2001.39.4.323.

Kim CH. Study on the Metagonimus sp. in Gum river basin, Chungchung-nam Do, Korea. *the Korean Journal of Parasitology.* 1980*, 18*: 215-228. doi:10.3347/kjp.1980.18.2.215.

Kim CH, & Guh JY. Study on Human Trematode Infections in the upper Basin of Gum River. *Biology Section Chungnam Journal of Sciences.* 1981*, 8*: 101-107.

Kim CH, Na YE, Kim NM, Shin DW, & Chang DY. Intestinal parasite and Clonorchis sinensis infection among the inhabitants in the upper stream of Taechong Dam, Kumgang (River). *the Korean Journal of Parasitology.* 1994*, 32*: 207-214. doi:10.3347/kjp.1994.32.4.207.

Kim CH, Park CH, Kim HJ, Chun HB, Min HK, Koh TY, & Soh CT. Prevalence Of Intestinal Parasites In Korea. *the Korean Journal of Parasitology.* 1971*, 9*: 25-38. doi:10.3347/kjp.1971.9.1.25.

Kim DC, Lee JS, Kim TS, Chang YM, Son SC, & Lee OY. Epidemiology of Clonorchiasis Endemicity of Clonorchis sinensis, in Sancheong area of Upper Nagdong River. *Report of NIH Korea.* 1984*, 54*: 267-286.

Kim GJ. *Reseach on status of helminthic Infection in chunnam: Centering on Gwagju city and sinan Kun*. 1983; Gwangju.

Kim IH. A Survey of Clonorchiasis in Korea. *Jeju Halla University Proceedings.* 1992*, 16*: 299-307.

Kim JH. *(A) studies on the status of helminthic infection of primary school students in urban and rural areas centered on Chungmu City and Kwangdo Myun, Tongyeong Gun*. 1984; Gyeongsangnam.

Kim SI, Jae G, & Park H. A Seroepidemiological Survey for Human Clonorchiasis on Soonchang-gun

Near the Sumjin River in Korea. *Korean Journal of Rural Medicine.* 2002*, 27*: 27-33.

Kim SI, Park J, Kim KS, Yang AH, & Kim YL. An Evaluation on the Prevalence and Reinfection after Medication of Patients with Clonorchis sinensis in an Endemic Locality. *Korean Journal of Rural Medicine.* 1999*, 24*: 225-232.

Kim SI, & Yun WS. Control of Human Clonorchiasis at Gokseong-gun and Sunchang-gun near the Sumjin River in Korea. *Korean Journal of Rural Medicine.* 2004*, 29*: 163-175.

Kim YH. Status of Intestinal Helminthes Infection in Primary School Children in Iksan, Korea. *Korean Journal of Clinical Laboratory Science.* 2007*, 39*: 86-90.

Kim YH, & Yu JP. Study on the Status of Helminthic Infections for Residents in the City Apartments. *Journal of public health & medical technology, Korea University.* 1975*, 6*: 47-51.

Lee BC, & Loh IK. An Epidemiological Survey on Helminthic Infections in a Rural Area. *Public Health Journal.* 1974*, 11*: 9-18.

Lee DM, Chung BJ, Chung JP, & Han DS. Prevalance of Clonorchis Sinensis and Nutritional Status of Children in an Endemic Area. *Korean Journal of Pediatrics.* 1975*, 18*: 20-24.

Lee GS. *Prevalence of clonorchiasis and its related factors among the inhabitants in Okcheon-gun, Korea*. 2008; Daejeon.

Lee GS, Cho IS, Lee YH, Noh HJ, Shin DW, Lee SG, & Lee TY. Epidemiological study of clonorchiasis and metagonimiasis along the Geum-gang (River) in Okcheon-gun (County), Korea. *the Korean Journal of Parasitology.* 2002*, 40*: 9-16. doi:10.3347/kjp.2002.40.1.9.

Lee JJ, Kim HJ, Kim MJ, Yi Lee JW, Jung BK, Lee JY, Shin EH, Kim JL, & Chai JY. Decrease of Metagonimus yokogawai Endemicity along the Tamjin River Basin. *the Korean Journal of Parasitology.* 2008*, 46*: 289-291. doi:10.3347/kjp.2008.46.4.289.

Lee JS, Joo KW, Chung YS, & Rim HJ. Parasitic Infection Among Ingabitants in Urban Area of Seoul. *Korean Journal of Rural Medicine.* 1985*, 10*: 36-41.

Lee JS, Lee WJ, Kho WG, Kim TS, In TS, Choi KS, & Song CY. Transition of endemicity of clonorchiasis in main riverside areas in Korea 1. Endemicity in Yechon and Sangju areas of upper Naktong-gang. *The Report National Institute of Health.* 1994a*, 31*: 172-182.

Lee JS, Lee WJ, Kho WG, Kim TS, In TS, Choi KS, & Song CY. Transition of endemicity of clonorchiasis in main riverside areas in Korea 2. Current status and changing pattern of the prevalence in the inhabitants in Kimhae areas of lower Naktong-gang. *The Report National Institute of Health.* 1994b*, 31*: 183-192.

Lee JS, Lee WJ, Kho WG, Lee HW, In TS, Yang YC, Kim DS, & Song CY. Serological study of Clonorchiasis. *The Report National Institute of Health.* 1995*, 32*: 188-200.

Lee JS, Lee WJ, Kim TS, In TS, Kim WS, & Kim SK. Current status and the changing pattern of the prevalence of clonorchiasis in the inhabitants in Sanchong-gun, Kyongsangnam-do, Korea. *the Korean Journal of Parasitology.* 1993*, 31*: 207-213. doi:10.3347/kjp.1993.31.3.207.

Lim MK, Ju YH, Franceschi S, Oh JK, Kong HJ, Hwang SS, Park SK, Cho SI, Sohn WM, Kim DI, Yoo KY, Hong ST, & Shin HR. Clonorchis sinensis infection and increasing risk of cholangiocarcinoma in the Republic of Korea. *American Journal of Tropical Medicine and Hygiene.* 2006*, 75*: 93-96. doi:10.4269/ajtmh.2006.75.93.

Min DY, Ryu JS, Ahn MH, Choi HK, Kang SI, & Shin MH. Present status of human paragonimiasis and intestinal parasitic infection in Bokildo (Islet), Korea. *Infection and Chemotherapy.* 2002*, 34*: 230-234.

Moon JG, Kim JJ, Moon CS, & Chung JH. A Study on the Status of Helminthic Infcetion in Urban and Rural Areas: Centering on Gwang-ju City and Bong-hwang Myun, Na-ju Gun. *The Medical Journal of Chosun University.* 1981*, 6*: 247-259.

Moon SL, Moon JG, & Chung YH. A Study on the Status of Hookworm Infection In Challanamdo. *The Medical Journal of Chosun University.* 1982*, 7*: 185-195.

Moon TG, Cho KK, & Chung YH. A survey on knowledge, attitudes and infection status of Parasites among housewives in Gwang Ju city. *The Medical Journal of Chosun University.* 1979*, 4*: 161-173.

Oh JK, Lim MK, Yun EH, Cho H, Park EY, Choi MH, Shin HR, & Hong ST. Control of clonorchiasis in Korea: effectiveness of health education for community leaders and individuals in an endemic area. *Tropical Medicine & International Health.* 2014*, 19*: 1096-1104. doi:10.1111/tmi.12338.

Park DS. Current Status of Clonorchis Sinensis Infection & Its Related Factors among the Residents of Rural Communities. *Korean Academy of Rural Health Nursing.* 2007*, 2*: 33-42. doi:10.22715/JKARHN.2007.2.1.033.

Park DS, Na SJ, Cho SH, June KJ, Cho YC, & Lee YH. Prevalence and Risk Factors of Clonorchiasis among Residents of Riverside Areas in Muju-gun, Jeollabuk-do, Korea. *the Korean Journal of Parasitology.* 2014*, 52*: 391-397. doi:10.3347/kjp.2014.52.4.391.

Park HY, Lee SU, Kim SH, Lee PC, Huh S, Yang YS, & Kong Y. Epidemiological significance of sero-positive inhabitants against sparganum in Kangwon-do, Korea. *Yonsei Medical Journal.* 2001*, 42*: 371-374. doi:10.3349/ymj.2001.42.4.371.

Park J, Kim KS, Ryu SY, Lee CG, Kim SI, Park H, Yang AH, & Kim YL. The prevalence of Clonorcihs sinensis and its associated factors at Goksung-gun area. *Korean Journal of Rural Medicine.* 2000*, 25*: 441-448.

Park JH, Guk SM, Shin EH, Kim HJ, Kim JL, Seo M, Park YK, & Chai JY. A new endemic focus of Gymnophalloides seoi infection on Aphae Island, Shinan-gun, Jeollanam-do. *the Korean Journal of Parasitology.* 2007*, 45*: 39-44. doi:10.3347/kjp.2007.45.1.39.

Park JH, Kim JL, Shin EH, Guk SM, Park YK, & Chai JY. A new endemic focus of Heterophyes nocens and other heterophyrid infections in a coastal area of Gangjin-gun, Jeollanam-do. *the Korean Journal of Parasitology.* 2007*, 45*: 33-38. doi:10.3347/kjp.2007.45.1.33.

Park OH. *Research on the Actual Condition of Parasitic Infections and Improvement of Preventive Management System in Gangwon-do*. 2008; Gangneung.

Park OS. *Study on the status of helminthic infections among Dongyeong middle school students and the riverside inhabitants in Cheongyoung Gun*. 1982; Daejeon.

Rhee JK, Baek BK, Lee SB, & Koh HB. Epidemiological studies of clonorchis sinensis in Mangyeong riverside area in Korea. *the Korean Journal of Parasitology.* 1983*, 21*: 157-166. doi:10.3347/kjp.1983.21.2.157.

Rim HJ, Joo KH, Eom KS, & Park SB. Epidemiological Note on the Clonorchiasis in Samrangjin Eup, Milyang Gun, Kyongsang Nam Do (=province). *Korean Journal of Rural Medicine.* 1982*, 7*: 80-89.

Rim HJ, Lee JS, Choi JO, Song OD, Song SD, & Kim MS. Epidemiological Survey on Clonorchis sinensis in Eui - Seong Gun, Kyungpook Province. *Korean Journal of Rural Medicine.* 1978*, 3*: 35-41.

Rim IS. *Epidemiological Studies on Clonorchiasis in the Area of Tam-Jin River*. 1985; Gwangju.

Ryu JC, Joo KH, Lee JS, & Rim HJ. Epidemiological Survey on Clonorchis sinensis Infection in Yedang Reservoir, Choong-cheong Namdo. *Korean Journal of Rural Medicine.* 1981*, 6*: 61-67.

Seo BS, Lee SH, Cho SY, Chai JY, Hong ST, Han IS, Sohn JS, Cho BH, Ahn SR, Lee SK, Chung SC, Kang KS, Shim HS, & Hwang IS. An Epidemiologic Study On Clonorchiasis And Metagonimiasis In Riverside Areas In Korea. *the Korean Journal of Parasitology.* 1981*, 19*: 137-150. doi:10.3347/kjp.1981.19.2.137.

Shen C, Kim JH, Lee JK, Bae YM, Choi MH, Oh JK, Lim MK, Shin HR, & Hong ST. Collection of Clonorchis sinensis adult worms from infected humans after praziquantel treatment. *the Korean Journal of Parasitology.* 2007*, 45*: 149-152. doi:10.3347/kjp.2007.45.2.149.

Shin DH, Kwak KW, & Joo CY. Epidemiological Studies of Clonorchis sinensis in the Vicinity of

Cheongdo River, Kyungpook, Korea. *Keimyung Medical Journal.* 1999*, 18*: 231-255.

Shin DW. A Parasitologial Survey in Chung Nam Area. *Chungnam medical journal.* 1977*, 4*: 129-136.

Shin HE, Lee MR, Ju JW, Jeong BS, Park MY, Lee KS, & Cho SH. Epidemiological and Clinical Parameters Features of Patients with Clonorchiasis in the Geum River Basin, Republic of Korea. *Interdisciplinary Perspectives on Infectious Diseases.* 2017*, 2017*: 7415301. doi:10.1155/2017/7415301.

Shin HR, Oh JK, Lim MK, Shin A, Kong HJ, Jung KW, Won YJ, Park S, Park SJ, & Hong ST. Descriptive epidemiology of cholangiocarcinoma and clonorchiasis in Korea. *J Korean Med Sci.* 2010*, 25*: 1011-1016. doi:10.3346/jkms.2010.25.7.1011.

Soh C-T, Min D-Y, Ryu J-S, & Yong T-S. Study on the Reproducibility of ELISA Technique for the Diagnosis of Clonorchiasis and Paragonimiasis. *YONSEI REPORTS ON TROPICAL MEDICINE.* 1985*, 16*: 1-10.

Soh CT, & Ahn YK. Epidemiological Study On Metagonimus Yokogawai Infection Along Boseong River In Jeonra Nam Do, Korea. *the Korean Journal of Parasitology.* 1978*, 16*: 1-13. doi:10.3347/kjp.1978.16.1.1.

Soh CT, Lee KT, Cho KM, Ahn YK, Kim SJ, Chung PR, Im KI, Min DY, Lee JH, & Chang JK. Prevalences of Clonorchiasis and Metagonimiasis along Rivers in Jeonra-Nam-Do, Korea. *YONSEI REPORTS ON TROPICAL MEDICINE.* 1976*, 7*: 3-16.

Sohn WM, Cho YP, Kim KJ, Kim MY, Lee CW, & Park DH. Analysis on the Findings of Serum Biochemical Test in Inhabitants Infected with Clonorchis sinensis. *Journal of biomedical laboratory sciences.* 2001*, 7*: 39-45.

Son WY, Huh S, Lee SU, Woo HC, & Hong SJ. Intestinal trematode infections in the villagers in Koje-myon, Kochang-gun, Kyongsangnam-do, Korea. *the Korean Journal of Parasitology.* 1994*, 32*: 149-155. doi:10.3347/kjp.1994.32.3.149.

Song HB, Kang MS, Park JS, Yoon SR, Choi BR, & Choi TY. *Distribution of parasites and pathogenic intestinal bacterial infections in Ebne-ri residents*. 1981; Jeonbuk.

Song IC, Lee JS, & Rim HJ. Epidemiological studies on the distribution of Clonorchis sinensis infection in Korea. *Korea University Medical School Magazine.* 1983*, 20*: 165-190.

Song SB. Epidemiological studies of Clonorchis sinensis in lower area of Nag Dong river nearby Busan City in Korea. *the Korean Journal of Parasitology.* 1982*, 20*: 133-141. doi:10.3347/kjp.1982.20.2.133.

Yu JR, Kwon SO, & Lee SH. Clonorchiasis and metagonimiasis in the inhabitants along Talchongang

(River), Chungwon-gun. *the Korean Journal of Parasitology.* 1994*, 32*: 267-269. doi:10.3347/kjp.1994.32.4.267.

Yun JI. *A Study on the infection rates of the Clonorchis sinensis in the residents and freshwater dishes around Chinju district*. 1991; Busan.

**Supplementary Table 2: Comparison of the prevalence of national survey and estimated prevalence**

| Year of survey | Prevalence in national survey (%) | Estimated prevalence (%, 95% BCI) |
|---|---|---|
| 1971 (***The Ministry of Health and Social Affairs and The Korean Association of parasite Eradication. 1971***) | 4.6 | 5.93 (4.76, 7.80) |
| 1976 (***The Ministry of Health and Social Affairs and The Korean Association of parasite Eradication. 1976***) | 1.8 | 4.82(4.05, 5.95) |
| 1981 (***The Ministry of Health and Social Affairs and The Korea Association of Parasite Eradication. 1982***) | 2.6 | 5.27(4.41, 6.38) |
| 1986 (***The Ministry of Health and Social Affairs and The Korea Association of Health. 1986***) | 2.7 | 2.40(2.01, 2.83) |
| 1992 (***The Ministry of Health and Social Affairs and The Korea Association of Health. 1993***) | 2.2 | 2.00(1.65, 2.45) |
| 1997 (***The Ministry of Health and Welfare and Korea Association of Health Promotion. 1997***) | 1.4 | 3.19(2.75, 3.73) |
| 2004 (***Korea Association of Health Promotion. 2004***) | 2.9 | 3.90(3.34, 4.51) |
| 2012 (***Korea Centers for Disease Control and Prevention and Korea National Institute of Health. 2013***) | 1.9 | 3.24(2.67, 3.89) |

**Reference**

The Ministry of Health and Social Affairs, The Korean Association of parasite Eradication. *Status of the first Korean intestinal parasite infection*. The Ministry of Health and Social Affairs, The Korean Association of parasite Eradication; 1971.

The Ministry of Health and Social Affairs, The Korean Association of parasite Eradication. *Prevalence of Intestinal Parasitic Infections in Korea-The 2nd Report*. The Ministry of Health and Social Affairs, The Korean Association of parasite Eradication; 1976.

The Ministry of Health and Social Affairs, The Korea Association of Parasite Eradication. *Prevalence of Intestinal Parasitic Infections in Korea-The Third Report*. The Ministry of Health and Social Affairs, The Korea Association of Parasite Eradication; 1982.

The Ministry of Health and Social Affairs, The Korea Association of Health. *Prevalence of Intestinal Parasitic Infections in Korea-The Fourth Report*. The Ministry of Health and Social Affairs, The Korea Association of Health; 1986.

The Ministry of Health and Social Affairs, The Korea Association of Health. *Prevalence of Intestinal Parasitic Infections in Korea-The Fifith Report*. The Ministry of Health and Social Affairs, The

Korea Association of Health; 1993.

The Ministry of Health and Welfare, Korea Association of Health Promotion. *Prevalence of intestinal parasitic infections in Korea-The Sixth Report.* The Ministry of Health and Welfare, Korea Association of Health Promotion; 1997.

Korea Association of Health Promotion. *Prevalence of Intestinal Parasitic Infections in Korea-The 7th Report*. Korea Association of Health Promotion; 2004.

Korea Centers for Disease Control and Prevention, Korea National Institute of Health. *National survey of the prevalence of Intestinal Parasitic Infections in Korea, the 8th Report*. Korea Centers for Disease Control and Prevention, Korea National Institute of Health; 2013.

**Supplementary Table 3: Posterior summaries of model parameters for *C. sinensis* infection by a sensitivity analysis**

| Variable | Estimated median with unadjusted diagnosis method[a] (95% BCI) | Estimated median with adjusted diagnosis method[b] (95% BCI) |
|---|---|---|
| Intercept | -5.41(-5.98, -4.85)[c] | -5.51(-6.07, -4.96)[c] |
| Survey type (Community)[d] | | |
| School | 1.46(1.20, 1.72)[c] | 1.54(1.27, 1.81)[c] |
| Urban extents (Rural)[d] | | |
| Urban | -0.53(-0.83, -0.24)[c] | -0.51(-0.8, -0.23)[c] |
| Land surface temperature in the daytime (≤17.0˚C)[d] | | |
| 17.0-18.5 | 0.13(-0.12, 0.39) | 0.14(-0.11, 0.39) |
| ＞18.5 | 0.24(-0.12, 0.61) | 0.24(-0.11, 0.6) |
| Annual precipitation (≤1266mm)[d] | | |
| 1263-1366 | -0.46(-0.82, -0.09)[c] | -0.45(-0.8, -0.1)[c] |
| ＞1366 | -0.51(-0.98, -0.04)[c] | -0.5(-0.95, -0.04)[c] |
| Elevation (≤56km)[d] | | |
| 56-129 | 0.04(-0.19, 0.28) | 0.04(-0.19, 0.27) |
| ＞129 | -0.18(-0.50, 0.15) | -0.17(-0.48, 0.14) |
| Distance to the nearest open water bodies (km) | -0.51(-0.65, -0.38)[c] | -0.5(-0.63, -0.37)[c] |
| Land surface temperature at night (°C) | 0.19(0.003, 0.39)[c] | 0.21(0.03, 0.4)[c] |
| Nighttime light | -0.06(-0.30, 0.18) | -0.07(-0.3, 0.15) |
| Normalized difference vegetation index | 0.24(0.06, 0.43)[c] | 0.24(0.06, 0.42)[c] |
| Range (km) | 45.60(38.60, 53.66) | 46.16(39.25, 54.35) |
| Spatial variance ($\sigma_{\phi}^2$) | 7.61(6.33, 9.19) | 6.95(5.75, 8.38) |
| Non-spatial variance ($\sigma^2_{nonsp}$) | 0.64(0.50, 0.81) | 0.59(0.46, 0.77) |
| Temporal correlation coefficient ($\rho$) | 0.04(-0.13, 0.20) | 0.05(-0.11, 0.21) |
| Variance of beta-likelihood ($\sigma^2_{\beta}$) | 0.01(0.01, 0.02) | 0.01(0.01, 0.02) |

[a]Estimated median with unadjusted diagnosis method: posterior parameter estimation with unadjusted diagnostic method in the final model.

[b]Estimated median with adjusted diagnosis method: posterior parameter estimation with adjusted diagnosis method.

[c]Important effect based on 95% Bayesian credible interval (BCI).

[d]In brackets, baseline values are reported.

**Supplementary Table 4: Posterior summaries of model parameters for *C. sinensis* infection by a sensitivity analysis**

| Variable | Estimated median in midpoint[a] (95% BCI) | Estimated median in low[b] (95% BCI) | Estimated median in upper[c] (95% BCI) |
|---|---|---|---|
| Intercept | -5.41(-5.98, -4.85)[d] | -5.38(-5.95, -4.82)[d] | -5.41(-5.96, -4.82)[d] |
| Survey type (Community)[e] | | | |
| School | 1.46(1.20, 1.72)[d] | 1.45(1.19, 1.72)[d] | 1.46(1.21, 1.73)[d] |
| Urban extents (Rural)[e] | | | |
| Urban | -0.53(-0.83, -0.24)[d] | -0.56(-0.87, -0.26)[d] | -0.52(-0.81, -0.23)[d] |
| Land surface temperature in the daytime (≤17.0˚C)[e] | | | |
| 17.0-18.5 | 0.13(-0.12, 0.39) | 0.11(-0.16, 0.37) | 0.15(-0.11, 0.40) |
| ＞18.5 | 0.24(-0.12, 0.61) | 0.22(-0.15, 0.60) | 0.26(-0.11, 0.63) |
| Annual precipitation (≤1266mm)[e] | | | |
| 1263-1366 | -0.46(-0.82, -0.09)[d] | -0.49(-0.86, -0.12)[d] | -0.45(-0.81, -0.08)[d] |
| ＞1366 | -0.51(-0.98, -0.04)[d] | -0.49(-0.97, -0.01)[d] | -0.50(-0.97, -0.03)[d] |
| Elevation (≤56km)[e] | | | |
| 56-129 | 0.04(-0.19, 0.28) | 0.04(-0.20, 0.28) | 0.03(-0.20, 0.26) |
| ＞129 | -0.18(-0.50, 0.15) | -0.19(-0.53, 0.14) | -0.17(-0.50, 0.15) |
| Distance to the nearest open water bodies (km) | -0.51(-0.65, -0.38)[d] | -0.51(-0.65, -0.37)[d] | -0.51(-0.64, -0.37)[d] |
| Land surface temperature at night (°C) | 0.19(0.003, 0.39)[d] | 0.24(0.04, 0.43)[d] | 0.18(-0.02, 0.37) |
| Nighttime light | -0.06(-0.30, 0.18) | -0.09(-0.33, 0.15) | -0.05(-0.29, 0.19) |
| Normalized difference vegetation index | 0.24(0.06, 0.43)[d] | 0.23(0.03, 0.42)[d] | 0.25(0.06, 0.44)[d] |
| Range (km) | 45.60(38.60, 53.66) | 44.16(37.61, 51.89) | 45.84(38.92,53.23) |
| Spatial variance ($\sigma_{\phi}^{2}$) | 7.61(6.33, 9.19) | 7.40(6.14, 8.94) | 7.62(6.32, 9.19) |
| Non-spatial variance ($\sigma^2_{nonsp}$) | 0.64(0.50, 0.81) | 0.69(0.55, 0.87) | 0.63(0.50, 0.79) |
| Temporal correlation coefficient ($\rho$) | 0.04(-0.13, 0.20) | 0.03(-0.14, 0.21) | 0.03(-0.19, 0.20) |
| Variance of beta-likelihood ($\sigma^2_{\beta}$) | 0.01(0.01, 0.02) | 0.06(0.04, 0.08) | 0.02(0.01, 0.02) |

[a]Estimated median in midpoint: posterior parameter estimation of the midpoint values of the intervals in the final model.

[b]Estimated median in upper: posterior parameter estimation of the upper limits of the prevalence intervals.

[c]Estimated median in low: posterior parameter estimation of the lower limits of the prevalence intervals.

[d]Important effect based on 95% Bayesian credible interval (BCI).

[e]In brackets, baseline values are reported.

**Supplementary Table 5: Remote sensing data sources for the current study related to in Korea[a]**

| Source | Data type | Data period | Temporal resolution | Spatial resolution |
|---|---|---|---|---|
| MODIS/Terra[b] | LST[c] | 2000-2018 | 8 days | 1km |
| MODIS/Terra[b] | NDVI[d] | 2000-2018 | 16 days | 1km |
| MODIS/Terra[b] | Land cover | 2001-2018 | Yearly | 1km |
| DIVA-GIS[e] | Elevation | 2000 | - | 1km |
| WorldClim[f] | Annual precipitation | 1960-1990 | - | 1km |
| DIVA-GIS[e] | Water bodies | 2000 | - | 30m |
| The Atlas of the Biosphere[g] | Soil moisture | 1950-1999 | - | 50km |
| Worldpop[h] | Population | 2000-2020 | - | 1km |
| SEDAC[i] | HII[j] | 1995-2004 | - | 1km |
| GHSL[k] | Urban extents | 1995 | - | 1km |
| NOAA[l] | Nighttime light | 1992-2008 | Yearly | 1km |
| Malaria atlas[m] | Travel time to the nearest big city | 2015 | - | 1km |
| UN[n] | Population growth rate | - | - | Country-level |

[a]Data asccessed in Janurary 2021

[b]Moderate Resolution Imageing Spectroradiometer (MODIS) /Terra, available at: https://lpdaac.usgs.gov/.

[c]Land surface temperature (LST) in the daytime and at night.

[d]NDVI: Normalized difference vegetation index.

[e]Available at: https://www.diva-gis.org/gdata/.

[f]Available at: http://www.worldclim.org/current/.

[g]Available at: http://www.sage.wisc.edu/atlas/.

[h]Available at: https://www.worldpop.org/geodata/.

[i]Socioeconomic Data and Applications Center, available at: http://sedac.ciesin.org/.

[j]HII: Human influence index.

[k]Global Human Settlement Layer, available at: http://data.jrc.ec.europa.eu/collection/GHSL/.

[l]National Ocranic And Atimospheric Administration, available at: https://www.ngdc.noaa.gov/.

[m]Available at:https://map.ox.ac.uk/research-project/accessibility_to_cities/

[n]UN: United Nations, available at: https://population.un.org/wpp/Download/Standard/Population/.

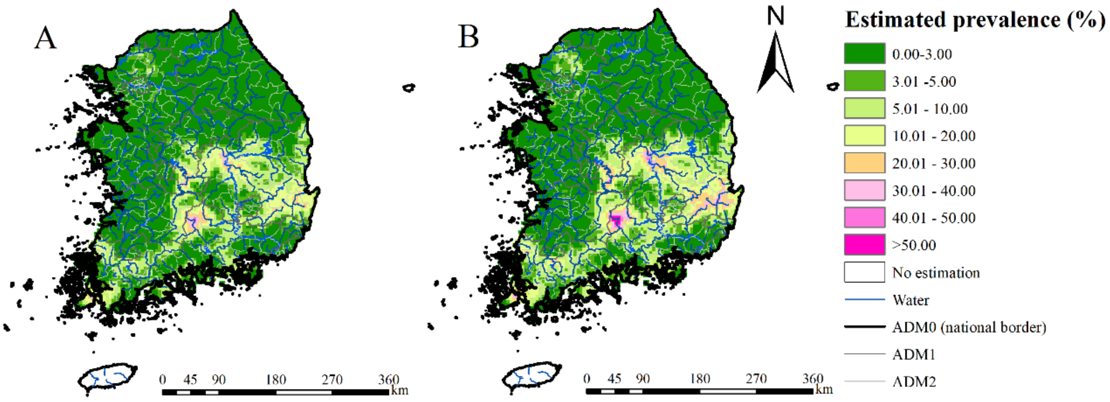

**Supplementary Figure 1: Model-based estimated risk maps of *C. sinensis* infection in 2017 for adjusted diagnosis method (A) and adjusted diagnosis method (B).**

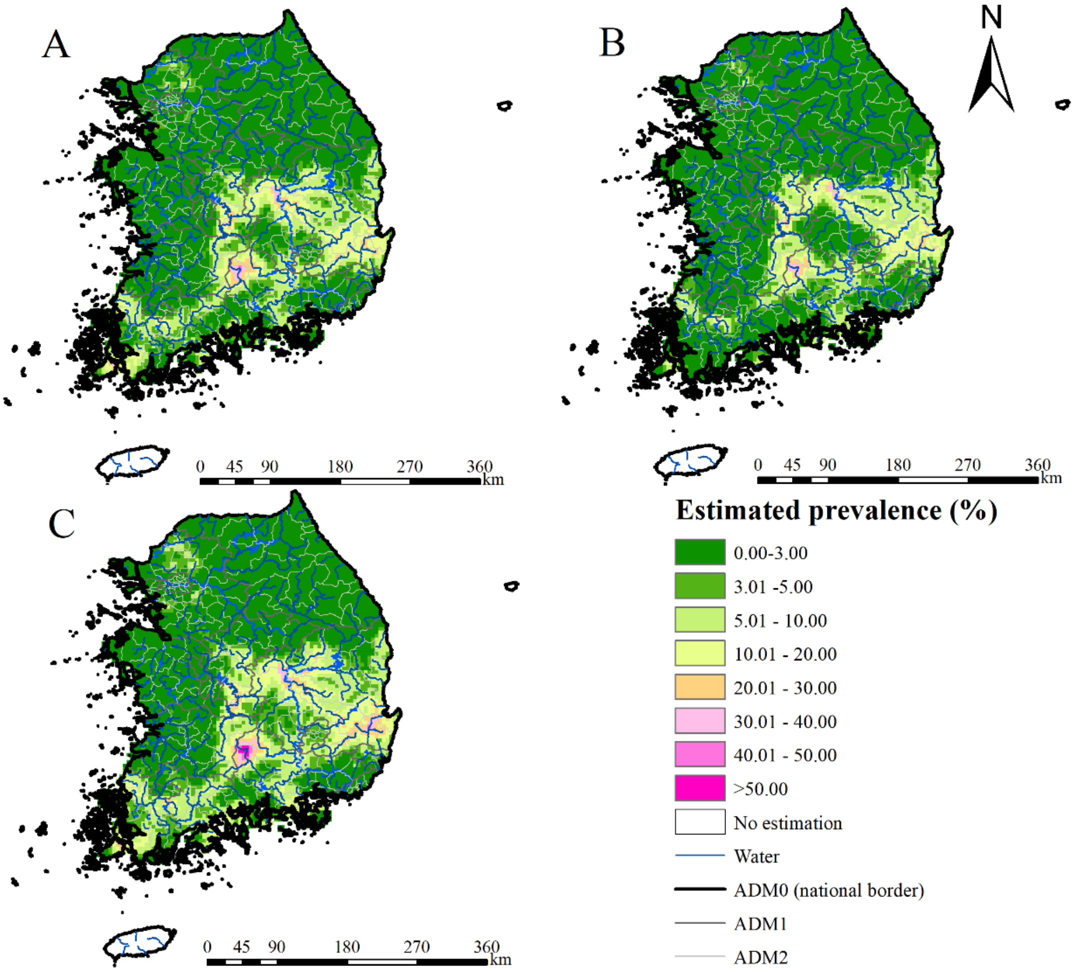

**Supplementary Figure 2: Model-based estimated risk maps of *C. sinensis* infection in 2017 under different values assigned to prevalence for surveys only reported prevalence in intervals.** (A) the median values, (B) the lower limits and (C) the upper limits of the intervals were assigned to prevalence.

**Literature search**

-Peer-review via two general databases (PubMed and ISI Web of Science) and three Korea databases (NAVER, KISS and RISS) with search terms "(Liver fluke* OR Clonorchi*) AND Korea" and ""간흡충" OR "간디스토마" OR "Clonorchis sinensis" OR Clonorchiasis", respectively. All references are merged in one reference database using EndNote X9.2 (Thomson Research Soft Ltd.).

-Google Scholar: general check for large disease database and other papers not included in the search engines.

↓

**Exclusion criteria**

(a) in-vitro investigations, or absence of human studies; (b) with specific study designs (e.g., case-control studies, intervention studies without prevalence at baseline or control group) or specific population groups (e.g., patients, migrant population) that could not represent the status of infection risk in corresponding study locations; (c) with survey locations where preventive chemotherapy treatment took place within one year; (d) with study locations/areas not clearly identified, or conducted in islands far away from the mainland; (e) with diagnostic methods including direct smear, salt water flotation, serum diagnostics, intradermal test, which are low sensitivity or difficult to differ the past and the active infection, or with diagnostic methods unidentified; (f) aggregated with community- and school-based surveys; and (g) with sample size less than 10.

↓

**Inclusion criteria**

prevalence related surveys (i.e., with information on number of examined and number of positive, or information on prevalence) conducted from 1970 onwards, reporting at point- or areal levels.

↓

**Identification of potentially relevant publications according to inclusion and exclusion criteria**

-Duplicates publications are checked and removed.

-Screen titles and abstracts to identify potential relevant articles.

-Full-text review to identify potentially relevant articles.

-During full text review, the potential relevant cited references of the articles are also screened to supplement the papers not collected earlier.

↓

**Quality control**

Quality control is undertaken by re-checking 20% of randomly selected irrelevant papers.

↓

**Geolocation**

The coordinates of the survey sites were obtained from the corresponding publications or from Google Maps (https://www.google.com/maps/) if they are not provided by the corresponding publications.

↓

**Prevalence data extraction**

-Detailed information of the selected literatures are extracted, including literature information (eg, journal, authors, publication date, title, volume and issue), survey information (eg, survey type, survey time), location information (eg, location names, location types, coordinates) and opisthorchiasis data (eg, diagnostic method, population type and age, numbers of examined, positive and percentage of positive).

-All extracted data are double-checked again to remove duplicates.

-For multiple papers with the same survey data, the data is extracted only once.

**Contact authors**

Authors of publications are contacted in case important information is missing (eg, prevalence, number of examined, number of positive, survey year).

**Quality assessment**

The quality assessment of each literature is undertaken using a nine-point checklist. The items of quality evaluation are as follows:

Q1: provide specific inclusion and exclusion criteria.

Q2: provide basic characteristics of the investigated population (gender, age, etc.).

Q3: provide prevalence rate of the survey.

Q4: provide number of positive patients and number of examined people of the survey.

Q5: provide diagnostic method used in the survey.

Q6: provide survey type.

Q7: provide time of the survey.

Q8: describe or discuss the possible bias of the survey or how confounders are controlled.

Q9: the literature comes from Science Citation Index Expanded database or Korea Citation Index

Each item is scored 1 in case the publication meets) or 0 in contrary. The scores are summed up for all items and assigned to the publication as its quality score.

**Data process and analysis**

-Data downloaded and process for environmental, socioeconomic data.

-Missing data imputation.

If survey year is missing, we assign year=public year minus three as survey year.

For surveys reported prevalence in intervals without exact observed values, the medians of the intervals were assigned.

-Geostatistical analysis.

A Bayesian geostatistical joint modeling approach is applied to analyze areal-level and point-referenced survey data together, including disease survey data reporting both the number of examined and positive, and those only reporting prevalence

Sensitivity analysis was conducted to assess the effect of using the midpoint values of intervals as the observed prevalence, and the effect of different diagnostical methods

**Supplementary Figure 3. Study protocol**

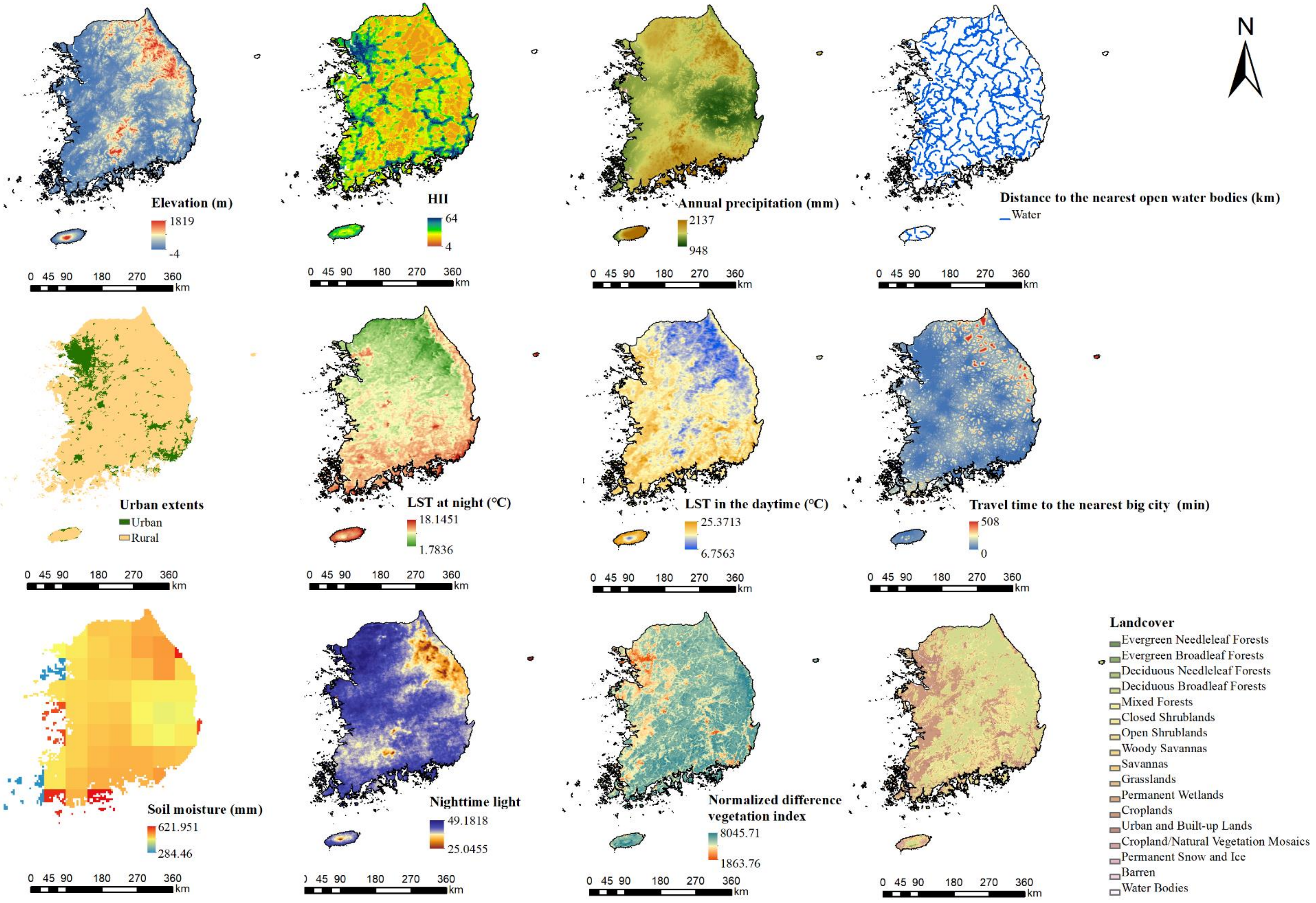

**Supplementary Figure 4: Images of spatial covariates used in the present study**
